# The burden of active infection and anti-SARS-CoV-2 IgG antibodies in the general population: Results from a statewide survey in Karnataka, India

**DOI:** 10.1101/2020.12.04.20243949

**Authors:** Giridhara R Babu, Rajesh Sundaresan, Siva Athreya, Jawaid Akhtar, Pankaj Kumar Pandey, Parimala S Maroor, MR Padma, R Lalitha, Mohammed Shariff, Lalitha Krishnappa, CN Manjunath, MK Sudarshan, G Gururaj, TS Ranganath, Vasanth Kumar, Pradeep Banandur, R Deepa, Shilpa Shiju, Eunice Lobo, Asish Satapathy, Lokesh Alahari, Prameela, T Vinitha, Anita Desai, V Ravi

**Affiliations:** Indian Institute of Public Health – Bengaluru, Public Health Foundation of India, Magadi Rd 1st cross, Next to leprosy hospital, SIHFW premises, Bengaluru, Karnataka 560023; Indian Institute of Science, CV Raman Rd, Bengaluru, Karnataka 560012; Indian Statistical Institute – Bangalore Centre, 8th Mile, Mysore Rd, RVCE Post, Bengaluru, Karnataka 560059; Department of health and family welfare services, Government of Karnataka, Vikasa soudha, Bengaluru, Karnataka 560008; Department of health and family welfare services, Aarogya Soudha, 1^st^ cross, Magadi road, Bengaluru, Karnataka 560023; Department of health and family welfare services Aarogya Soudha, 1^st^ cross, Magadi road, Bengaluru, Karnataka 560023; State maternal and PPTCT consultant, UNICEF, Bengaluru; M S Ramaiah Medical College, M S Ramaiah Nagar, Mathikere, Bengaluru, Karnataka 560054; Sri Jayadeva Institute of Cardiovascular Sciences and Research, Bannerghatta Main Rd, Phase 3, Jayanagara 9th Block, Jayanagar, Bengaluru, Karnataka 560069; Technical Advisory Committee on COVID19, Department of health and family welfare services Aarogya Soudha, 1^st^ cross, Magadi road, Bengaluru, Karnataka 560023; National Institute of Mental Health and Neurosciences, Hosur Road, Bengaluru, Karnataka; Bangalore Medical College and Research Institute, Fort, K.R. Road, Bengaluru, 560002; National Institute of Mental Health and Neurosciences. Bengaluru, Karnataka; Indian Institute of Public Health-Bengaluru, Public Health Foundation of India, Magadi Rd 1st cross, Next to leprosy hospital, SIHFW premises, Bengaluru, Karnataka 560023; WHO – NPSP, Member Technical Advisory Committee on COVID19, Bengaluru; National Institute of Mental Health and Neurosciences, Hosur Road, Bengaluru, Karnataka 560029

## Abstract

**Background:** Globally, the routinely used case-based reporting and IgG serosurveys underestimate the actual prevalence of COVID-19. Simultaneous estimation of IgG antibodies and active SARS-CoV-2 markers can provide a more accurate estimation.

**Methods:** A cross-sectional survey of 16416 people covering all risk groups was done between 3-16 September 2020 using the state of Karnataka’s infrastructure of 290 hospitals across all 30 districts. All participants were subjected to simultaneous detection of SARS-CoV-2 IgG using a commercial ELISA kit, SARS-CoV-2 antigen using a rapid antigen detection test (RAT), and reverse transcription-polymerase chain reaction (RT-PCR) for RNA detection. Maximum-likelihood estimation was used for joint estimation of the adjusted IgG, active, and total prevalence, while multinomial regression identified predictors.

**Findings:** The overall adjusted prevalence of COVID-19 in Karnataka was 27 ·3% (95% CI: 25 ·7-28 ·9), including IgG 16 ·4% (95% CI: 15 ·1 - 17 ·7) and active infection 12 ·7% (95% CI: 11 ·5-13 ·9). The case-to-infection ratio was 1:40, and the infection fatality rate was 0 ·05%. Influenza-like symptoms or contact with a COVID-19 positive patient are good predictors of active infection. The RAT kits had higher sensitivity (68%) in symptomatic participants compared to 47% in asymptomatic.

**Interpretation:** This is the first comprehensive survey providing accurate estimates of the COVID-19 burden anywhere in the world. Further, our findings provide a reasonable approximation of population immunity threshold levels. Using the RAT kits and following the syndromic approach can be useful in screening and monitoring COVID-19. Leveraging existing surveillance platforms, coupled with appropriate methods and sampling framework, renders our model replicable in other settings.

## INTRODUCTION

The global pandemic of SARS-CoV-2 causing coronavirus disease (COVID-19) has raged across the world within a few months. India has the second-highest burden of COVID-19 with 8 ·8 million infected and 130070 deaths, as of 16 November 2020.^1^ Currently, India has only case-based reporting as the prime strategy through all the epidemic phases. Case-based reporting has the advantages of rationalizing testing, isolating cases, and tracing and quarantining contacts.^2^ However, it does not provide an estimate of the true burden of the disease, as it picks up mostly sicker people seeking care or those who have better access to health care. Hence, the reported case counts of COVID-19 grossly underestimate the true prevalence of the pandemic. The two rounds of national seroprevalence surveys conducted by the Indian Council of Medical Research (ICMR) indicated that for every reported case, 81-130 infections were missed in the initial survey conducted in May 2020,^3^ which improved to missing nearly 26–32 infections per reported case by August 2020. This may be underestimated as it captured only IgG prevalence, and the sampling was not representative. Serological surveys, such as those conducted by the ICMR, can help understand the burden of past infections. However, detecting new cases is challenging since 45% of the infected people have mild or no symptoms.^4^ Furthermore, the estimation of active infections is affected by poor in-person testing due to inaccessibility, stigma, and supply-side inadequacies.

Effective public health measures require understanding the existing burden of disease reliably through epidemiological investigations. Joint estimation of IgG prevalence and active SARS-CoV-2 infections can help detect, manage, and control the disease outbreak. Seroprevalence estimates from the world show varying numbers ranging from 0 ·07% in-hospital patients to 54 ·1% in slum inhabitants.^5-13^ Although not representative, the ICMR survey results reported 0 ·73% prevalence across the country (May-June 2020), which increased to 7% by the end of September.^3^ The surveys in slums and non-slums of Mumbai^5^ showed considerable variation, 54 ·1% (95% CI: 52 ·7 to 55 ·6) and 16 ·1% (95% CI: 14 ·9 to 17 ·4) prevalence, respectively. Serosurveys in a healthcare setting of North India showed prevalence increasing from 2 ·3% in April to 50 ·6% in July.^14^ However, there are concerns about using only IgG prevalence as a marker of population immunity threshold. These include the inability to detect the IgG antibodies over time, varying sampling methods, the unreliable nature of the predictive value of positive antibody tests with varying sensitivity and specificity of different tests affecting the tests’ reliability, and the presence of other types of the immune response. ^15-17^ This can be resolved to a great extent by understanding the burden of active infections concurrently along with IgG estimation.

Karnataka has an estimated population of 70 ·7 million spread over 191791 square kilometers. The first confirmed COVID-19 case was reported on 09 March 2020. As of 16 November 2020, there were 861,647 cumulative cases, 27,146 active cases, and 11,529 deaths.^1^ Our goal was to estimate the IgG prevalence and active prevalence of SARS-CoV-2 infections in Karnataka jointly and assess the variation across geographical regions and risk groups. Here we report the results of what is perhaps the first comprehensive statewide survey in India. The design and analysis methodologies of this survey can serve as the blueprint for other similar surveys.

## METHODS

### Study setting, design, and sample size

#### Setting

This was the first round of the proposed serial cross-sectional surveys across the districts of Karnataka. This state has 30 administrative districts. The capital district Bengaluru has approximately 13 ·6 million residents. The study was conducted during 03-16 September 2020.

#### Design

Each district was a unit of the survey except Bengaluru, which was subdivided into 9 units. From the resulting geographically representative 38 units, health facilities with the expertise to conduct the survey were selected (Figure 1 and Appendix D). The participants included only adults aged 18 years and above. The survey excluded those already diagnosed with SARS-CoV-2 Infection, those unwilling to provide a sample for the test or those who did not agree to provide informed consent. The population was stratified into three risk groups based on community exposure and vulnerability to COVID-19. The Low-risk group comprised pregnant women presenting for antenatal check-up (ANC) clinic, and persons attending the outpatient department for common ailments in the hospitals and their attendees. The Moderate-risk group comprised persons with high contact in the community, bus conductors and autorickshaw drivers, vendors at vegetable markets, healthcare workers, individuals in containment zones, persons in congregate settings (markets, malls, retail stores, bus stops, railway stations), and waste collectors.^18^ The High-risk group comprised the elderly (60 years of age and above) and persons with co-morbid conditions (chronic liver disease, chronic lung disease, chronic renal disease, diabetes, heart disease, hypertension, immunocompromised condition, malignancy).

**Figure 1:**
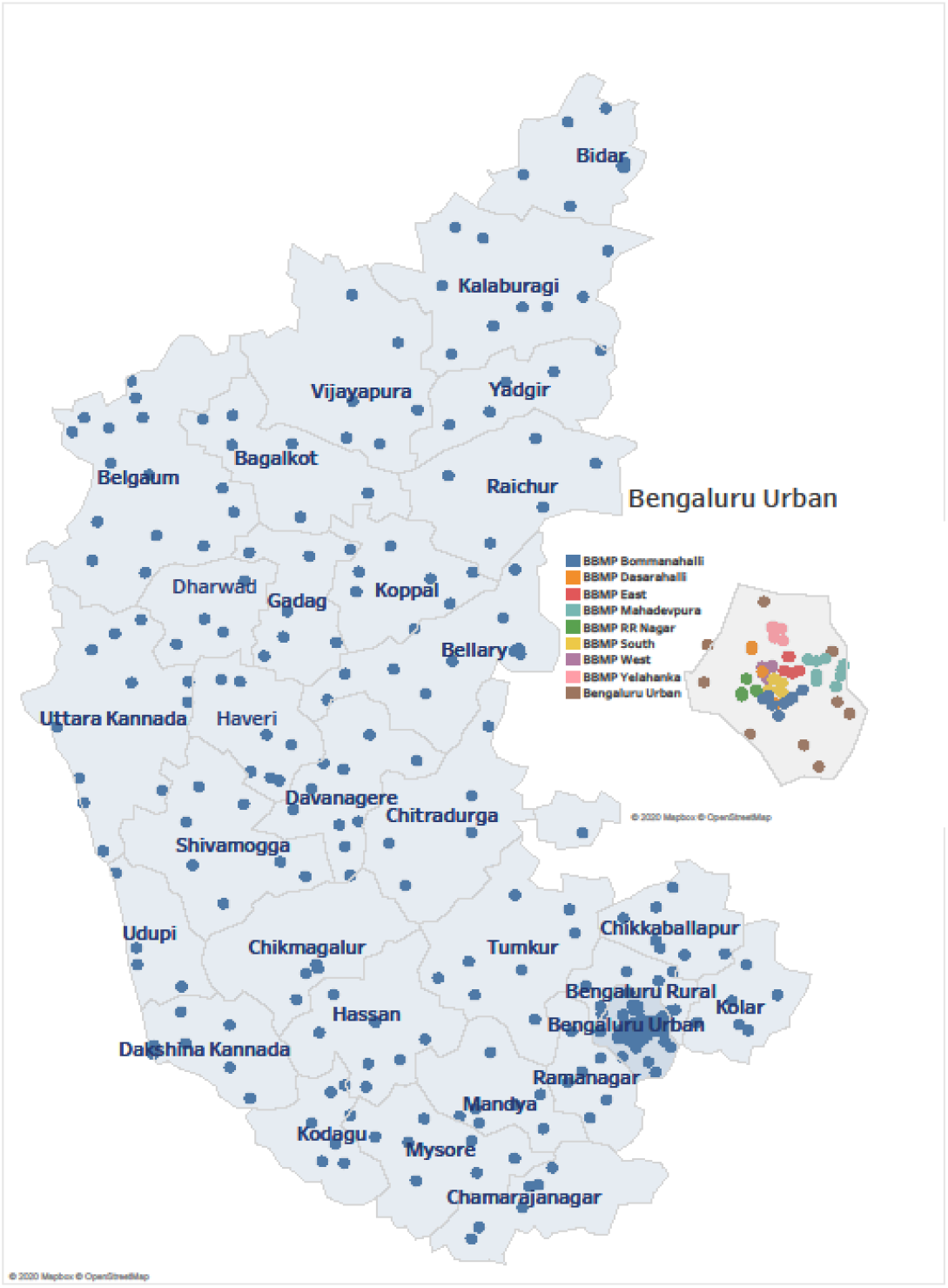
Sites (blue dots) of the survey representing geographical spread across Karnataka. The inset picture shows the sites across Bengaluru (multi-colored dots).

#### Sample size

Assuming 10% prevalence, the minimum sample size of 432 per cluster, for a target 95% confidence level, a margin of error 0 ·05, and design effect 3, led to a total sample size of 16416 across the 38 units.

### Sample collection and laboratory testing

From participants in the low-risk group (Figure 2), we collected both nasopharyngeal and oropharyngeal swab samples for the RT-PCR test following the ICMR protocol and 4 ml of venous blood for the IgG antibody test. In the moderate and high-risk groups, we collected two swab samples in different media for the antigen and RT-PCR tests and 4 ml of venous blood.

**Figure 2:**
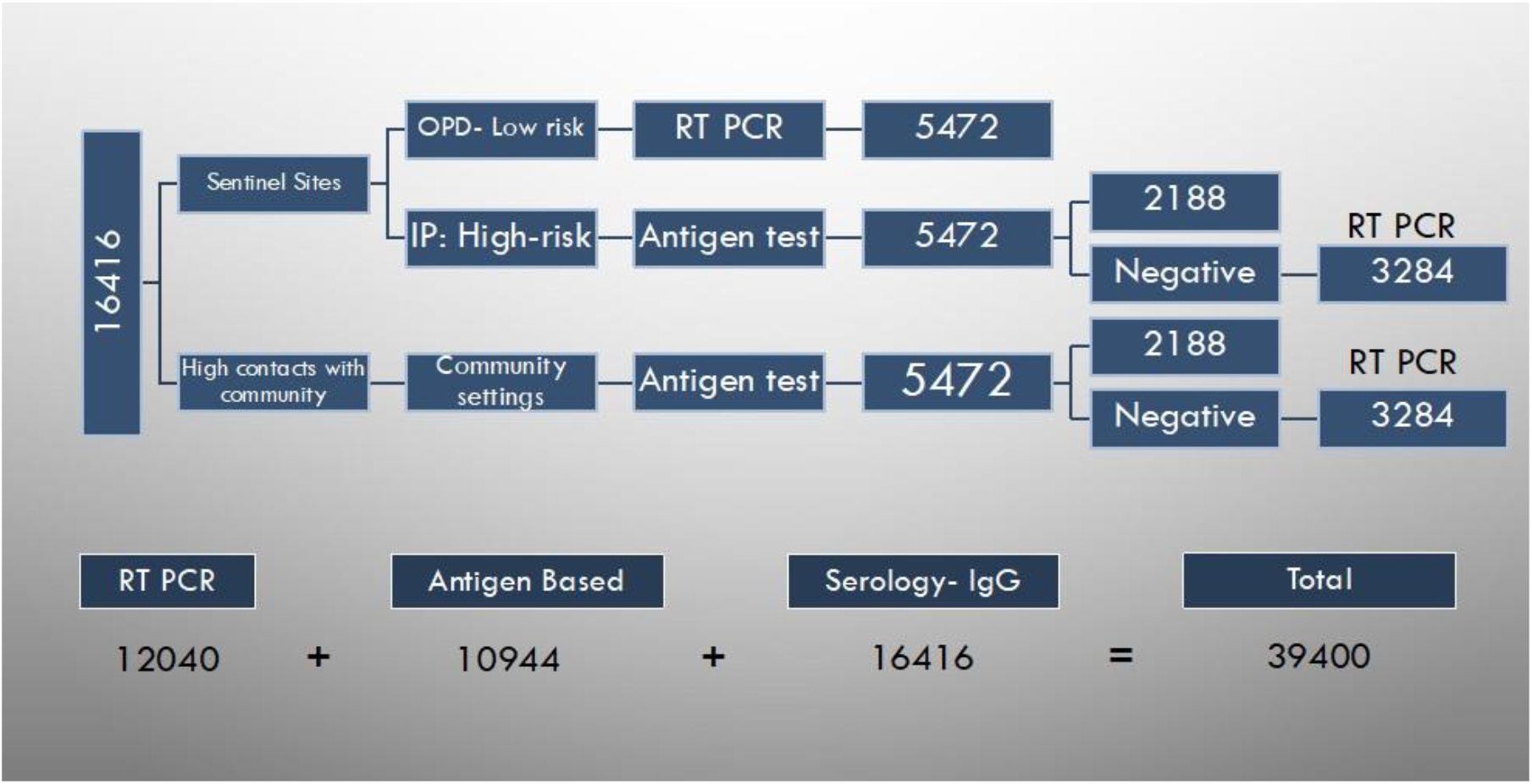
Schema for different tests in the survey, Round 1, Karnataka state. A total of 16,416 participants assigned to 38 units across the state targeted 432 participants per unit. Within the units, the 432 were divided into 144 participants from each risk-group – low risk, moderate risk, and high risk. Only the RT-PCR and antibody tests were conducted for the low risk group. All three tests were conducted for the moderate and high risk groups. In the event RAT being positive, the RT-PCR tests were not conducted for the participants. The estimated number of RT-PCR tests, Antigen tests, and IgG antibody tests were 12040, 10944, and 16416, respectively.

The rapid antigen detection test (RAT) was done using the *Antigen Standard Q COVID-19 Ag detection kit*, a rapid chromatographic immunoassay for the qualitative detection of antigens specific to SARS-CoV-2. The RT-PCR test was done on all low-risk participants and on those who tested negative on the RAT (Figure 3) through the current ICMR-approved testing network. For antibody testing, the collected venous blood sample was left undisturbed at room temperature for 30 minutes for clotting, then centrifuged at 3000 rpm, and the serum was transported to the laboratory by maintaining a cold chain. SARS-CoV-2-specific IgG antibodies were detected using a commercially available, validated, and ICMR-approved kit (Covid Kavach Anti SARS-CoV-2 IgG antibody detection ELISA, Zydus Cadila, India)^19^. The test was performed as per the manufacturer’s instructions. The results were interpreted as positive or negative for SARS-CoV-2 IgG antibodies based on the cut-off value of optical densities obtained with positive and negative samples provided in the kit.

**Figure 3:**
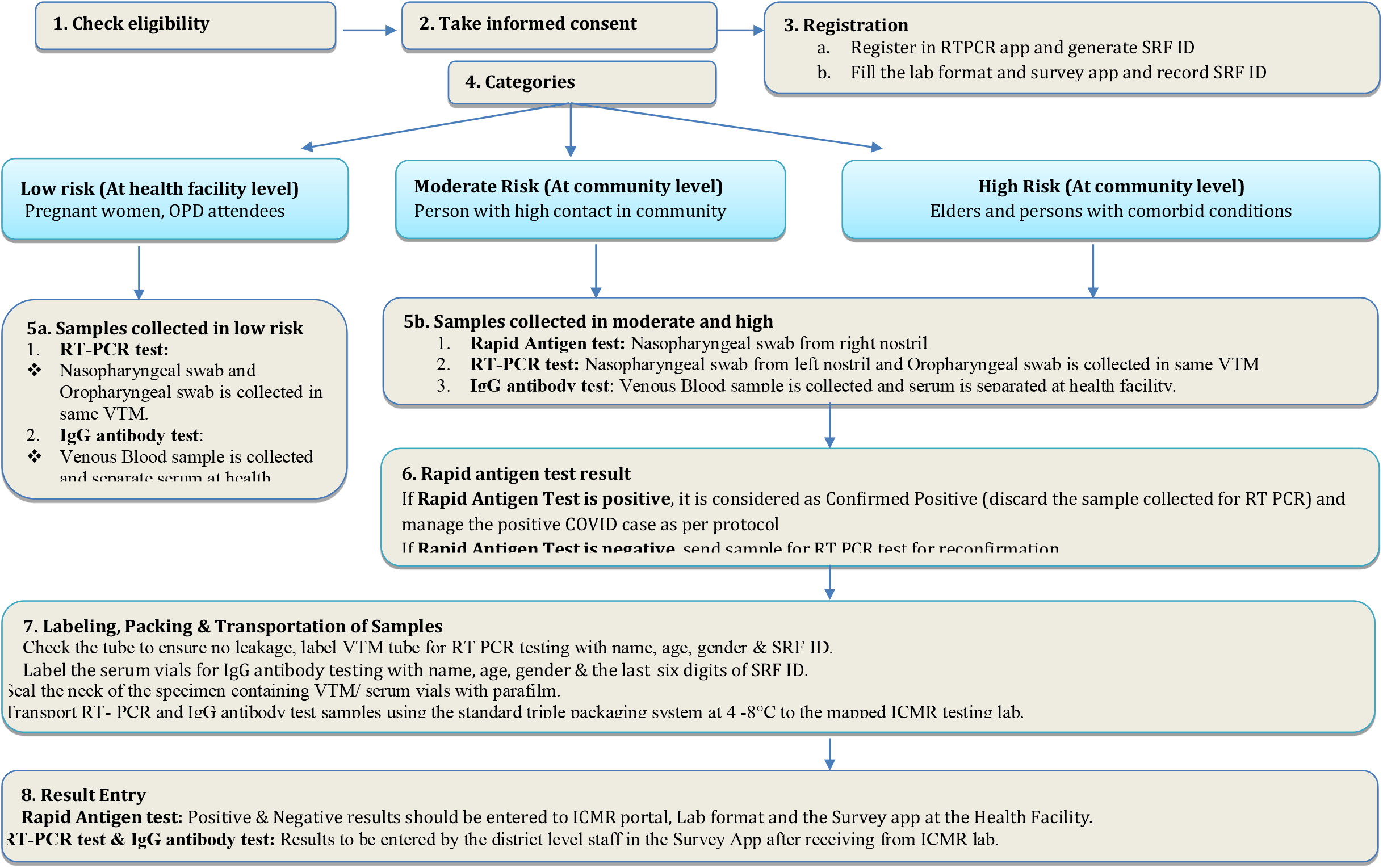
Algorithm for the serial cross-sectional survey for estimating COVID-19 burden in the Karnataka state. Step-1 checks for exclusion criteria, Step-2 is obtaining consent, Step-3 indicates the registration procedure. A participant is automatically categorized as low risk, moderate risk or high risk. The protocol and the procedure for the survey are detailed in the rest of the flow chart.

### Data collection

After obtaining written informed consent, information on basic demographic details, exposure history to laboratory-confirmed COVID-19 cases, symptoms suggestive of COVID-19 in the preceding one month, and clinical history were recorded on a web-based application designed specifically for the study and were linked to the samples using the ICMR Specimen Referral Forms for COVID-19. The category, symptoms, contact, and comorbidity information for participants were gathered using the web-based application. RAT/RT-PCR results were entered into the ICMR test-data portal. The IgG antibody test results were retrieved directly from the labs. A consolidated line-list of all the participants was then created. From this, subsets of participants in risk-categories, subcategories, age groups, sex, and geographical units were used to jointly estimate IgG prevalence, active infection, and total burden in the respective categories. Symptoms and comorbidity data, which were part of the consolidated line-list, were used in the regression. Only anonymized data with no personal identifiers were used for the analysis.

### Ethical considerations

The Institutional Ethics Committee (IEC) of the Indian Institute of Public Health-Bengaluru campus reviewed and approved the study (vide. IIPHHB/TRCIEC/174/2020). Participants’ test results were available and shared with them by the concerned health facility.

### Statistical analysis

To enable *joint estimation* of IgG prevalence, active infection, and total prevalence, we first modelled an individual to be in one of four disease states: having active infection but no IgG antibodies, having IgG antibodies but no evidence of active infection, having both IgG antibodies and active infection, and finally having neither active infection nor IgG antibodies. The disease state of the individual is, however, *hidden* and can only be inferred from the RAT, the RT-PCR, and the IgG antibody test outcomes. This leads to a *parametric model* for the probabilities of test outcomes (observations) given the disease-state probabilities (parameters of the model), after taking the sensitivities and the specificities of the tests into account.

To get the joint estimates of the parameters in a stratum, we use *maximum likelihood estimation*, which *ipso facto* provides estimates already *adjusted* for the sensitivities and the specificities of the tests. The joint estimation is an extension of the Rogan-Gladen formula.^20^ The procedure also accounts for the protocol-induced variation of test-types across participants. *Confidence intervals* are obtained by invoking asymptotic normality of the maximum likelihood estimates, with their covariance matrix being approximated by the inverse of the *Fisher information* matrix of the parametric model. Further, *weighted adjusted estimates* for Karnataka were obtained after weighing each district’s prevalence estimates by the population fraction in that district. We computed *odds ratios* by restricting attention to the relevant subcategories.

To identify the weights on various independent/explanatory variables (symptoms, comorbidities, etc.) for predicting past infection and active infection, we use *multinomial regression* to regress the test outcomes on the independent variables. The procedure can be embedded within the framework of the *generalized linear model* with multinomial logit functions along with a *custom link function* that accounts for not only the test-type variability across participants but also the tests’ sensitivities and specificities. Important explanatory variables are captured using the *Wald test*. The details are given in the supplementary material provided.

## RESULTS

Of the 16585 people surveyed in the different risk categories, we present the results for 15624 individuals whose RAT plus RT-PCR and *COVID Kavach ELISA* antibody test results have been line-matched (Appendix C). A total of 16585 IgG results were provided. The results of 513 were not considered due to missing information and the inability to match the participant in the database; 448 entries were further not mapped to the line-list because of manual data-entry errors or because data was not retrievable from the ICMR portal. Also, 18 IgG samples were inconclusive (Figure 1 in Supplementary Material/Appendix C).

### IgG prevalence

The overall weighted adjusted seroprevalence of IgG is 16 ·4% (95% CI: 15 ·1 – 17 ·7). This was as of 03 September 2020 and at the state level, obtained after adjusting for the serial sensitivities and specificities of all tests (Table 1).

**TABLE 1:**
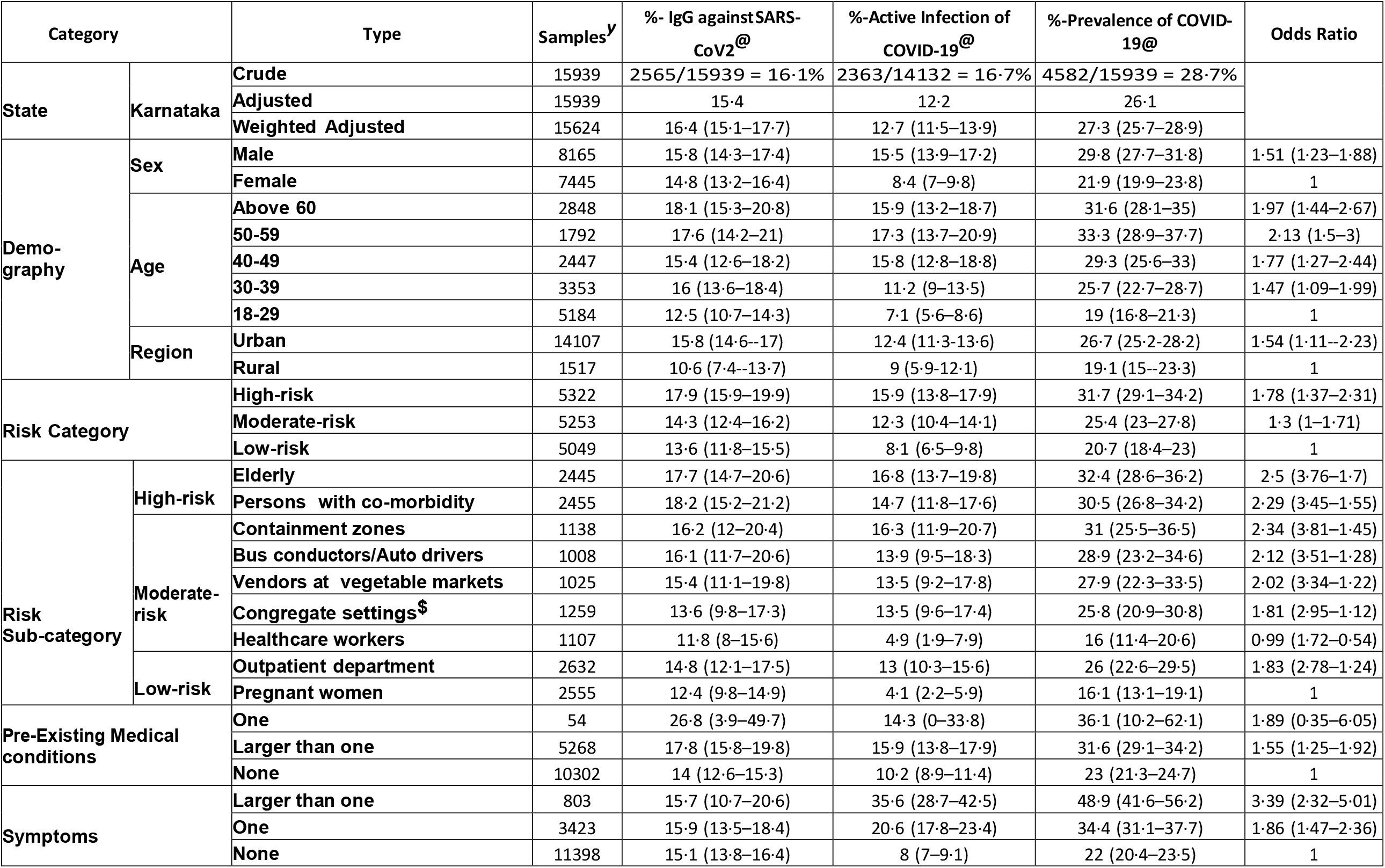
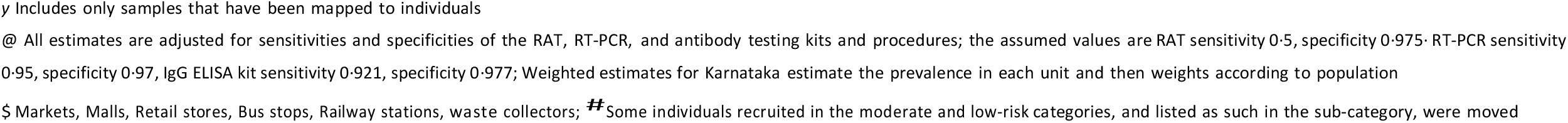
Seroprevalence of IgG antibodies against SARS-CoV2 and Active Infection in Karnataka.

| Category |  | Type | Samples<sup>y</sup> | %- IgG againstSARS-CoV2<sup>@</sup> | %-Active Infection of COVID-19<sup>@</sup> | %-Prevalence of COVID-19<sup>@</sup> | Odds Ratio |
| --- | --- | --- | --- | --- | --- | --- | --- |
| State | Karnataka | Crude | 15939 | 2565/15939 = 16.1% | 2363/14132 = 16.7% | 4582/15939 = 28.7% |  |
|  |  | Adjusted | 15939 | 15.4 | 12.2 | 26.1 |  |
|  |  | Weighted Adjusted | 15624 | 16.4 (15.1–17.7) | 12.7 (11.5–13.9) | 27.3 (25.7–28.9) |  |
| Demo-graphy | Sex | Male | 8165 | 15.8 (14.3–17.4) | 15.5 (13.9–17.2) | 29.8 (27.7–31.8) | 1.51 (1.23–1.88) |
|  |  | Female | 7445 | 14.8 (13.2–16.4) | 8.4 (7–9.8) | 21.9 (19.9–23.8) | 1 |
|  | Age | Above 60 | 2848 | 18.1 (15.3–20.8) | 15.9 (13.2–18.7) | 31.6 (28.1–35) | 1.97 (1.44–2.67) |
|  |  | 50-59 | 1792 | 17.6 (14.2–21) | 17.3 (13.7–20.9) | 33.3 (28.9–37.7) | 2.13 (1.5–3) |
|  |  | 40-49 | 2447 | 15.4 (12.6–18.2) | 15.8 (12.8–18.8) | 29.3 (25.6–33) | 1.77 (1.27–2.44) |
|  |  | 30-39 | 3353 | 16 (13.6–18.4) | 11.2 (9–13.5) | 25.7 (22.7–28.7) | 1.47 (1.09–1.99) |
|  |  | 18-29 | 5184 | 12.5 (10.7–14.3) | 7.1 (5.6–8.6) | 19 (16.8–21.3) | 1 |
|  | Region | Urban | 14107 | 15.8 (14.6--17) | 12.4 (11.3-13.6) | 26.7 (25.2-28.2) | 1.54 (1.11–2.23) |
|  |  | Rural | 1517 | 10.6 (7.4--13.7) | 9 (5.9-12.1) | 19.1 (15--23.3) | 1 |
| Risk Category |  | High-risk | 5322 | 17.9 (15.9–19.9) | 15.9 (13.8–17.9) | 31.7 (29.1–34.2) | 1.78 (1.37–2.31) |
|  |  | Moderate-risk | 5253 | 14.3 (12.4–16.2) | 12.3 (10.4–14.1) | 25.4 (23–27.8) | 1.3 (1–1.71) |
|  |  | Low-risk | 5049 | 13.6 (11.8–15.5) | 8.1 (6.5–9.8) | 20.7 (18.4–23) | 1 |
| Risk Sub-category | High-risk | Elderly | 2445 | 17.7 (14.7–20.6) | 16.8 (13.7–19.8) | 32.4 (28.6–36.2) | 2.5 (3.76–1.7) |
|  |  | Persons with co-morbidity | 2455 | 18.2 (15.2–21.2) | 14.7 (11.8–17.6) | 30.5 (26.8–34.2) | 2.29 (3.45–1.55) |
|  | Moderate-risk | Containment zones | 1138 | 16.2 (12–20.4) | 16.3 (11.9–20.7) | 31 (25.5–36.5) | 2.34 (3.81–1.45) |
|  |  | Bus conductors/Auto drivers | 1008 | 16.1 (11.7–20.6) | 13.9 (9.5–18.3) | 28.9 (23.2–34.6) | 2.12 (3.51–1.28) |
|  |  | Vendors at vegetable markets | 1025 | 15.4 (11.1–19.8) | 13.5 (9.2–17.8) | 27.9 (22.3–33.5) | 2.02 (3.34–1.22) |
| | | Congregate settings<sup>\$</sup> | 1259 | 13.6 (9.8–17.3) | 13.5 (9.6–17.4) | 25.8 (20.9–30.8) | 1.81 (2.95–1.12) |
|  |  | Healthcare workers | 1107 | 11.8 (8–15.6) | 4.9 (1.9–7.9) | 16 (11.4–20.6) | 0.99 (1.72–0.54) |
|  | Low-risk | Outpatient department | 2632 | 14.8 (12.1–17.5) | 13 (10.3–15.6) | 26 (22.6–29.5) | 1.83 (2.78–1.24) |
|  |  | Pregnant women | 2555 | 12.4 (9.8–14.9) | 4.1 (2.2–5.9) | 16.1 (13.1–19.1) | 1 |
| Pre-Existing Medical conditions |  | One | 54 | 26.8 (3.9–49.7) | 14.3 (0–33.8) | 36.1 (10.2–62.1) | 1.89 (0.35–6.05) |
|  |  | Larger than one | 5268 | 17.8 (15.8–19.8) | 15.9 (13.8–17.9) | 31.6 (29.1–34.2) | 1.55 (1.25–1.92) |
|  |  | None | 10302 | 14 (12.6–15.3) | 10.2 (8.9–11.4) | 23 (21.3–24.7) | 1 |
| Symptoms |  | Larger than one | 803 | 15.7 (10.7–20.6) | 35.6 (28.7–42.5) | 48.9 (41.6–56.2) | 3.39 (2.32–5.01) |
|  |  | One | 3423 | 15.9 (13.5–18.4) | 20.6 (17.8–23.4) | 34.4 (31.1–37.7) | 1.86 (1.47–2.36) |
|  |  | None | 11398 | 15.1 (13.8–16.4) | 8 (7–9.1) | 22 (20.4–23.5) | 1 |

y Includes only samples that have been mapped to individuals
@ All estimates are adjusted for sensitivities and specificities of the RAT, RT-PCR, and antibody testing kits and procedures; the assumed values are RAT sensitivity 0.5, specificity 0.975. RT-PCR sensitivity 0.95, specificity 0.97, IgG ELISA kit sensitivity 0.921, specificity 0.977; Weighted estimates for Karnataka estimate the prevalence in each unit and then weights according to population
\$ Markets, Malls, Retail stores, Bus stops, Railway stations, waste collectors; ~~##~~ Some individuals recruited in the moderate and low-risk categories, and listed as such in the sub-category, were moved

### Active infection

We estimate that 12 ·7% (95% CI: 11 ·5—13 ·9) of the seemingly unsuspected participants in the general population, or an estimated 89,88,313 (95% CI: 81,39,023—98,37,602) people, were having active infection (as on 16 September 2020). This is based on the numbers that tested positive on RT-PCR/RAT and after taking into account the IgG outcomes and the serial sensitivities and specificities of all tests (Table 1).

### Overall COVID-19 prevalence

The overall adjusted prevalence of COVID-19 at the state level was 27 ·3% (95% CI: 25 ·7 – 28 ·9) as of 16 September 2020 (combined IgG and active infection (Table 1)).

### Stratifications

The seroprevalence of IgG among males and females were similar, but the active infection was higher in males than females (15 ·5% vs · 8 ·4%) (Table 1, Figure 4). Thus, the overall prevalence was higher in males than in females (29 ·8% vs. 21 ·9%). Estimates of both seroprevalence and total prevalence were higher in the elderly population and low among the less-than-30-years-old population (Figure 5). The high-risk population had a higher prevalence (31 ·7% (CI: 29 ·1 – 34 ·2)), followed by the moderate-risk population (25 ·4% (CI: 23 ·0 – 27 ·8)) and then the low-risk population (20 ·7 (CI: 18 ·4 – 23 ·0)) (Table 1 and Figure 4).

**Figure 4:**
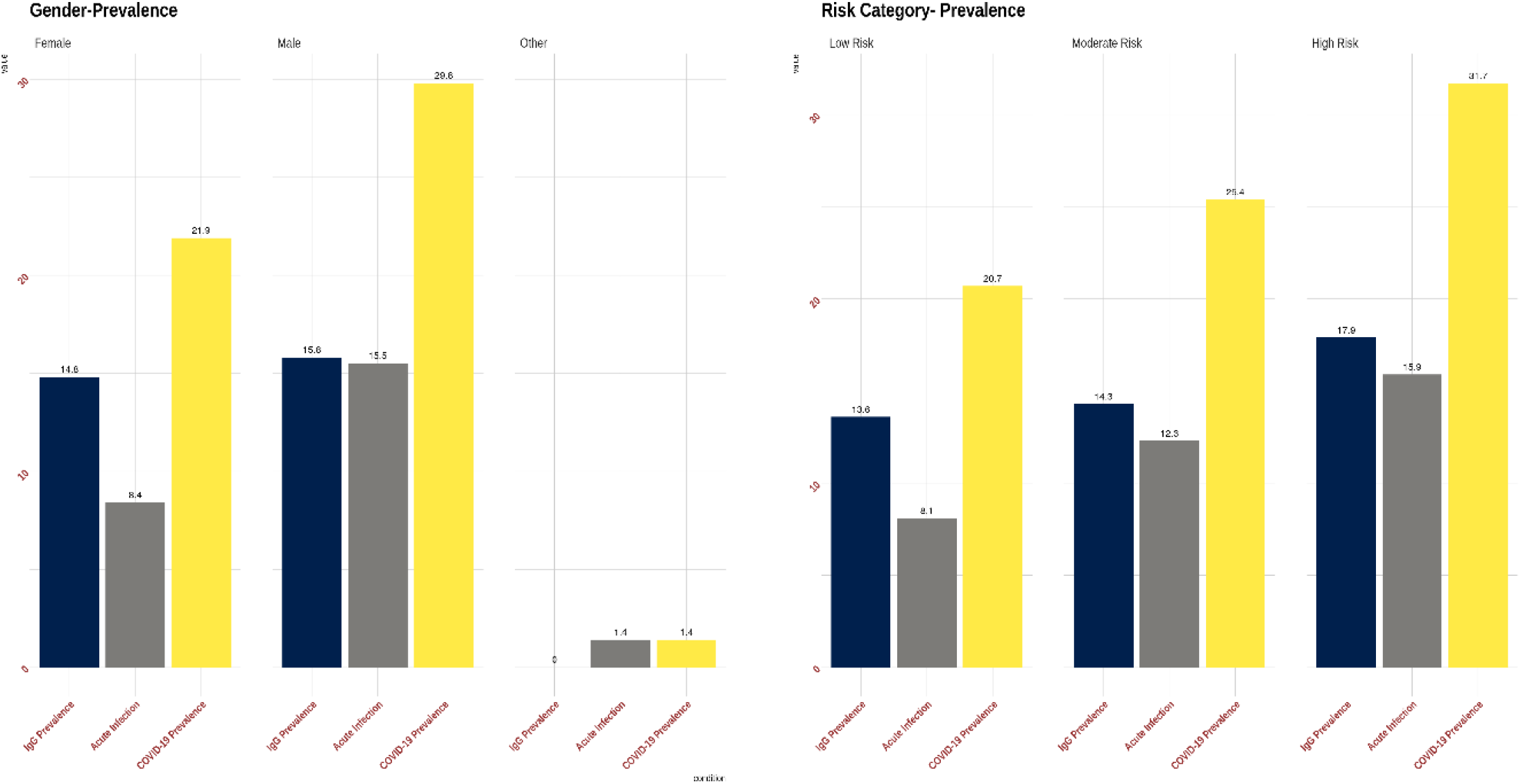
Prevalence in the state categorized according to sex and risk groups.

**Figure 5:**
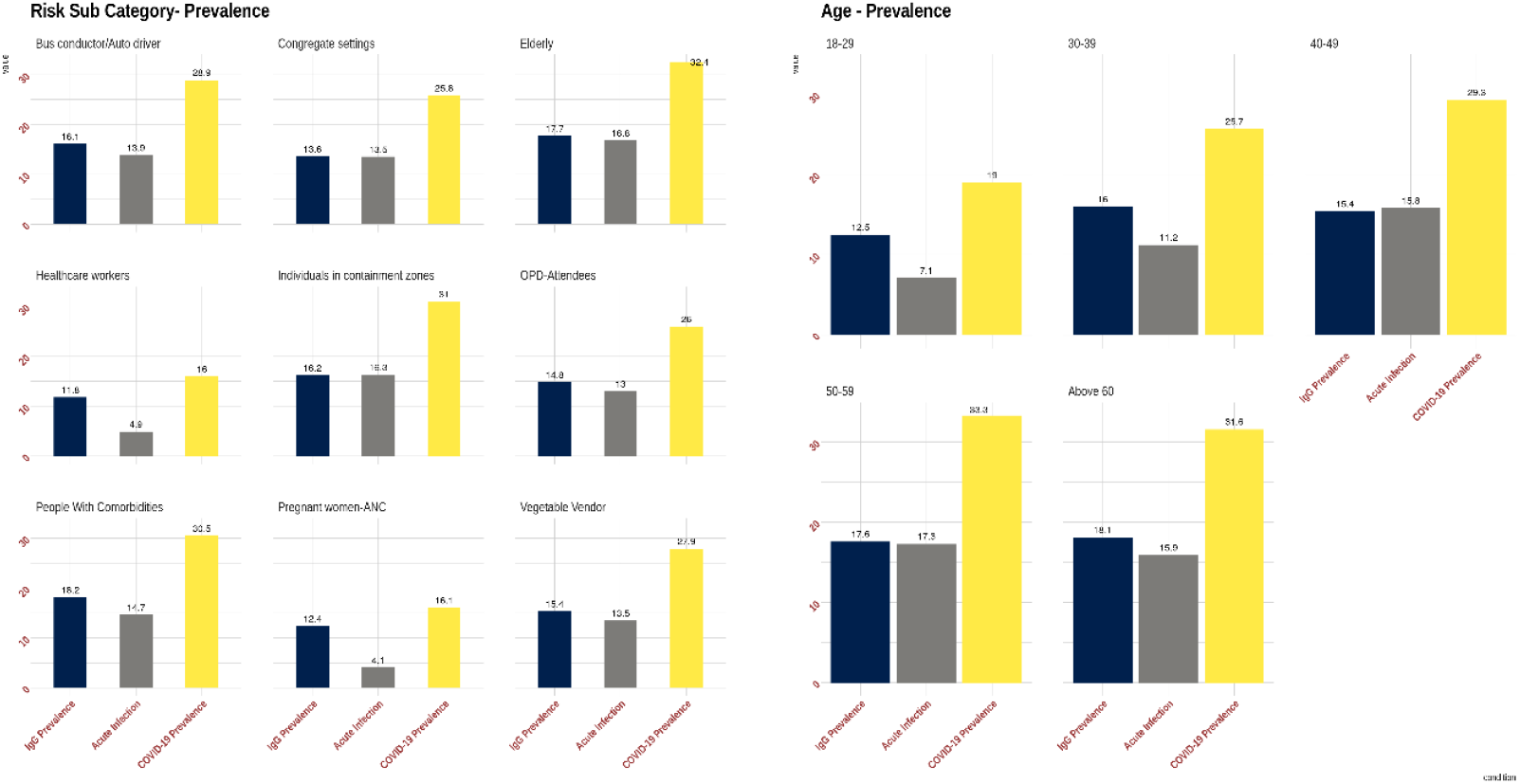
Prevalence (IgG, acute, total) across risk-subcategories and age groups in the state. The panel on the left depicts IgG seroprevalence (black bars) and active infection rates (Grey and yellow bars) based on risk categories, while the panel on the right depicts age-wise break up of seroprevalence rates.

### Case-to-infection ratio (CIR)

At the state level, it is estimated that for every RT-PCR confirmed case detected, there were 40 undetected infected individuals as of 16 September 2020 (Table 3 and Figure 6). This is estimated by using 484,954 reported number of cases in Karnataka^1^ and the adjusted prevalence of COVID-19 (27 ·3%) against SARS-CoV-2. The cases-to-infections ratio ranges from 10 to 111 across units.

**TABLE 3:** Unit wise Case-to-Infection Ratio (CIR) and Infection Fatality Rate (IFR) in Karnataka.

| Unit | Cases up to<br>16 September 2020 | Estimated<br>Infection | CIR | IFR |
| --- | --- | --- | --- | --- |
| Dharwad | 14927 | 182654 | 1:12 | 0.23% |
| BBMP Mahadevpura | 13906 | 150294 | 1:11 | 0.13% |
| Gadag | 7800 | 104140 | 1:13 | 0.12% |
| BBMP Bommanahalli | 18137 | 313581 | 1:17 | 0.11% |
| Dakshina Kannada | 18558 | 630499 | 1:34 | 0.11% |
| BBMP RR Nagar | 13147 | 262335 | 1:20 | 0.10% |
| Bengaluru Urban | 25294 | 250886 | 1:10 | 0.10% |
| Bagalkot | 8547 | 257014 | 1:30 | 0.09% |
| BBMP Yelahanka | 11559 | 120182 | 1:10 | 0.09% |
| Shivamogga | 13097 | 398975 | 1:30 | 0.09% |
| BBMP South | 28614 | 912400 | 1:32 | 0.08% |
| Hassan | 12850 | 564948 | 1:44 | 0.08% |
| Tumakuru | 10002 | 818525 | 1:82 | 0.08% |
| BBMP Dasarahalli | 8529 | 104054 | 1:12 | 0.07% |
| BBMP East | 27530 | 538197 | 1:20 | 0.07% |
| BBMP West | 33567 | 695302 | 1:21 | 0.07% |
| Bengaluru Urban Conglomerate | 180283 | 4060572 | 1:23 | 0.07% |
| Chikkaballapur | 6125 | 165082 | 1:27 | 0.07% |
| Mysuru | 27486 | 917989 | 1:33 | 0.07% |
| Chikmagalur | 6704 | 361425 | 1:54 | 0.06% |
| Davanagere | 13840 | 853580 | 1:62 | 0.06% |
| Karnataka | 484954 | 19321334 | 1:40 | 0.05% |
| Udupi | 14278 | 475836 | 1:33 | 0.05% |
| Ballari | 27512 | 1334597 | 1:49 | 0.04% |
| Bidar | 5597 | 355963 | 1:64 | 0.04% |
| Haveri | 7062 | 503099 | 1:71 | 0.04% |
| Kalaburagi | 14979 | 886977 | 1:59 | 0.04% |
| Kolar | 4709 | 273049 | 1:58 | 0.04% |
| Koppal | 9478 | 355495 | 1:38 | 0.04% |
| Uttara Kannada | 7633 | 247149 | 1:32 | 0.04% |
| Kodagu | 2086 | 114963 | 1:55 | 0.03% |
| Belgaum | 17043 | 1611769 | 1:95 | 0.02% |
| Bengaluru Rural | 7165 | 327092 | 1:46 | 0.02% |
| Chamarajanagar | 3179 | 227054 | 1:71 | 0.02% |
| Mandya | 8752 | 468436 | 01:54 | 0.02% |
| Raichur | 9821 | 745933 | 1:76 | 0.02% |
| Ramanagar | 4471 | 334188 | 1:75 | 0.02% |
| Vijayapura | 8172 | 910433 | 1:111 | 0.02% |
| Yadgir | 7312 | 446333 | 1:61 | 0.02% |
| Chitradurga | 5486 | 467031 | 1:85 | 0.01% |

**Figure 6:**
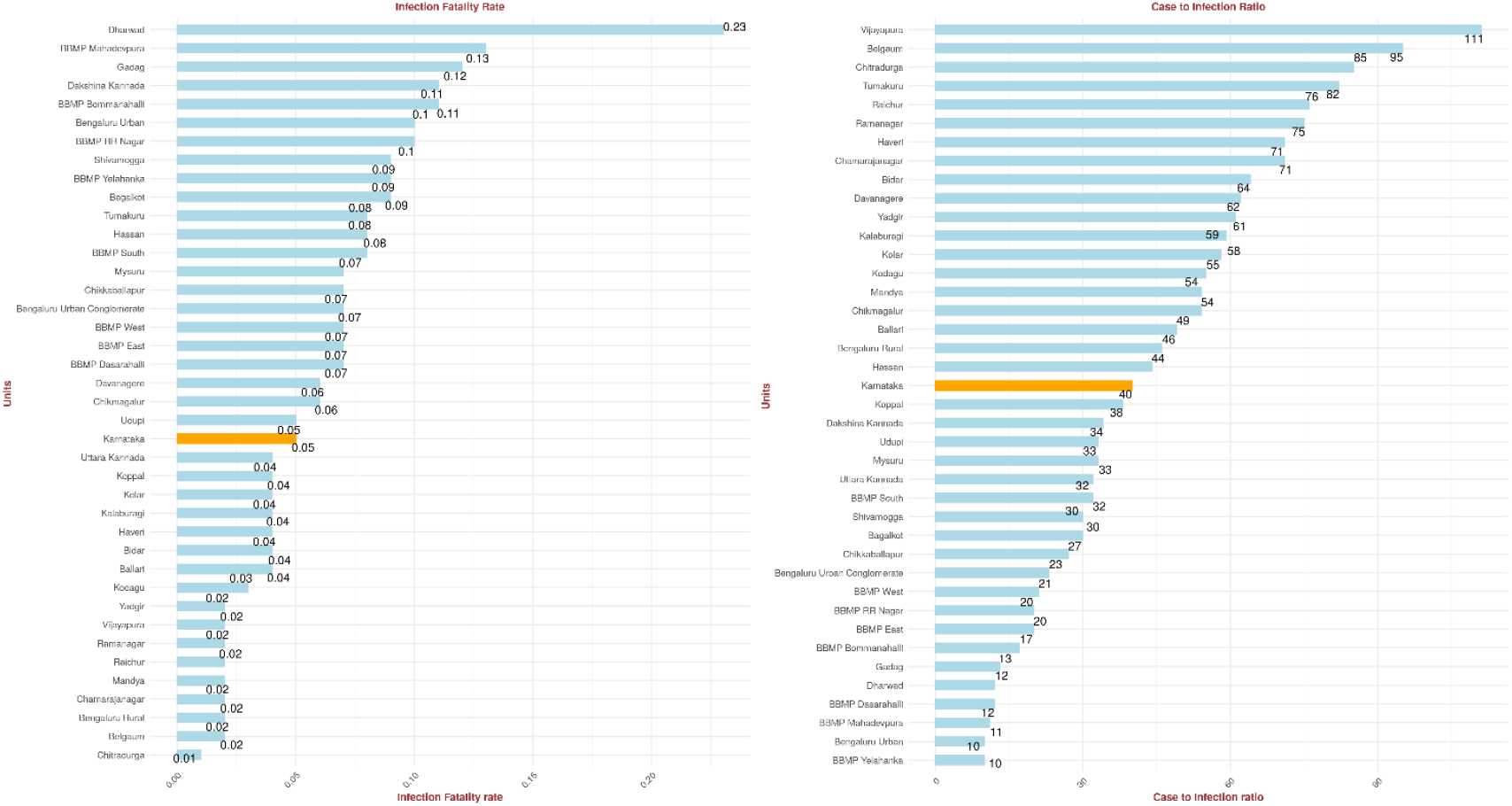
Infection-fatality-rate and Case-to-infection ratio across the different units. Each sub-graph is ordered according to the values. The orange bar represents the value for the entire state of Karnataka.

### Infection fatality rate (IFR)

As of 03 September 2020, the IFR due to COVID-19 in Karnataka is estimated as 0 ·05%, with more than half the units (21 out of 38) above state IFR. The highest was estimated in the Dharwad district (0 ·23%) (Table 3 and Figure 6).

### District/unit variations across the state

The IgG prevalence was highest in Vijayapura district (23 ·9%) and lowest in Bagalkot district (4 ·1%). The state capital Bengaluru had an IgG prevalence of 22% (95% CI: 19 ·1 – 24 ·9). The active infection was highest in Ballari (34 ·5%) and lowest in Bidar (0 ·7%). Bengaluru’s active infection was an estimated 9 ·2% (95% CI: 7 ·1 - 11 ·3). The overall COVID-19 prevalence was lowest in Dharwad district (8 ·7%) and highest in Ballari district (43 ·1%) (Table 2, Figures 7 and 8). The overall COVID-19 prevalence in Bengaluru was estimated to be 29 ·8% (95% CI: 26 ·5 −33). Within Bengaluru itself (with *N* = 3617 samples), we estimated that BBMP West had the highest IgG against SARS-CoV-2 and prevalence of COVID-19. In contrast, BBMP Mahadevapura had the least (Figures 7 and 9, and Supplementary Table 1). Again, BBMP RR Nagar had the highest active infection within Bengaluru, and BBMP East had the lowest (Figure 7, Supplementary material Table 1). Districts with high cases infections ratio (more than 40) are Vijayapura, Belgaum, Chitradurga, Tumakuru, Raichur, Ramanagar, Haveri, Chamarajanagar, Bidar, Davanagere, Yadgir, Kalaburagi, Kolar, Kodagu, Mandya, Chikmagalur, Ballari, Bengaluru Rural, Hassan (Table 3 and Figure 6). To summarize in a sentence, there is differential exposure to the disease across the state (Figure 8).

**TABLE 2:** Seroprevalence of IgG antibodies against SARS-CoV2 and Active Infection in districts of Karnataka state (*N* = 15624)

| Unit | Samples<sup>y</sup> | %- IgG against SARS-CoV2<sup>†</sup> | %-Active Infection<sup>†</sup> | %-Prevalence of COVID-19<sup>†</sup> |
| --- | --- | --- | --- | --- |
| **Karnataka** | 15624 | 16.4 (15.1–17.7) | 12.7 (11.5–13.9) | 27.3 (25.7–28.9) |
| **Ballari** | 406 | 22.1 (14.3–29.9) | 34.5 (25.4–43.6) | 43.1 (33.5–52.6) |
| **Davanagere** | 412 | 16.4 (9.4–23.4) | 29.2 (20.3–38.1) | 40.6 (31–50.3) |
| **Udupi** | 439 | 16.2 (9.5–23) | 22.8 (15.1–30.5) | 36.4 (27.5–45.4) |
| **Vijayapura** | 381 | 23.9 (15.7–32.2) | 13.9 (6.6–21.1) | 35.4 (25.7–45.1) |
| **Raichur** | 404 | 22.8 (14.9–30.7) | 12.1 (5.5–18.7) | 34.1 (24.7–43.4) |
| **Chikmagalur** | 436 | 12 (5.9–18.1) | 21 (13.2–28.8) | 31.8 (22.8–40.8) |
| **Yadgir** | 422 | 15.4 (8.6–22.1) | 18.6 (11.2–26) | 31.6 (22.7–40.5) |
| **Hassan** | 410 | 13.2 (6.7–19.7) | 21.2 (12.9–29.5) | 30.7 (21.3–40) |
| **Belgaum** | 430 | 23.7 (16–31.5) | 6.4 (1.4–11.5) | 30.1 (21.4–38.9) |
| **Kalaburagi** | 425 | 17.1 (10.1–24.1) | 14.5 (7.8–21.1) | 29.8 (21.1–38.4) |
| **Bengaluru Urban Conglomerate** | 3617 | 22 (19.1–24.9) | 9.2 (7.1–11.3) | 29.8 (26.5–33) |
| **Tumakuru** | 429 | 6.8 (1.7–11.8) | 25.2 (16.2–34.2) | 29.4 (19.9–38.9) |
| **Ramanagar** | 408 | 13.9 (7.2–20.6) | 16.2 (8.7–23.6) | 29.3 (20.2–38.5) |
| **Bengaluru Rural** | 432 | 15.2 (8.6–21.9) | 16.5 (9–23.9) | 28.7 (19.8–37.6) |
| **Haveri** | 417 | 14.8 (8.1–21.5) | 14.6 (7.8–21.4) | 28.6 (19.9–37.4) |
| **Mysuru** | 402 | 18.8 (11.4–26.2) | 8.4 (2.7–14.1) | 27.2 (18.4–36) |
| **Dakshina Kannada** | 430 | 14.5 (8–21.1) | 13.5 (7–20.1) | 27 (18.5–35.5) |
| **Chitradurga** | 411 | 10.2 (4.2–16.1) | 16 (8.5–23.4) | 25.9 (17–34.8) |
| **Mandya** | 414 | 18.5 (11.2–25.9) | 6.7 (1.3–12.2) | 25.3 (16.6–33.9) |
| **Koppal** | 427 | 19.6 (12.3–26.9) | 2.7 (0–6.2) | 22.3 (14.3–30.2) |
| **Shivamogga** | 426 | 7.7 (2.4–13) | 13.7 (6.8–20.6) | 21.4 (13.1–29.7) |
| **Chamarajanagar** | 383 | 15.8 (8.6–22.9) | 6.6 (1.1–12.1) | 21.1 (12.7–29.5) |
| **Kodagu** | 412 | 12 (5.8–18.3) | 8.7 (2.8–14.6) | 20.5 (12.4–28.7) |
| **Bidar** | 407 | 18 (10.7–25.2) | 0.7 (0–3.3) | 18.7 (11–26.3) |
| **Uttara Kannada** | 419 | 8.1 (2.6–13.5) | 8.7 (3–14.4) | 16.3 (8.8–23.8) |
| **Kolar** | 431 | 10.1 (4.3–15.9) | 6.8 (1.6–11.9) | 16.1 (8.8–23.5) |
| **Chikkaballapur** | 412 | 6.4 (1.3–11.5) | 5.9 (0–11.8) | 12.1 (4.5–19.7) |
| **Bagalkot** | 401 | 4.1 (0–8.6) | 9.7 (3.6–15.8) | 12 (5–19.1) |
| **Gadag** | 341 | 6.3 (0.8–11.8) | 2.7 (0–8.5) | 9 (1.1–17) |
| **Dharwad** | 440 | 7.1 (2–12.1) | 2 (0–5.6) | 8.7 (2.7–14.7) |

<sup>y</sup> Includes only samples that have been mapped to individuals
<sup>†</sup> Adjusted for sensitivities and specificities of RAT, RT-PCR, and antibody testing kits and procedure

**Figure 7:**
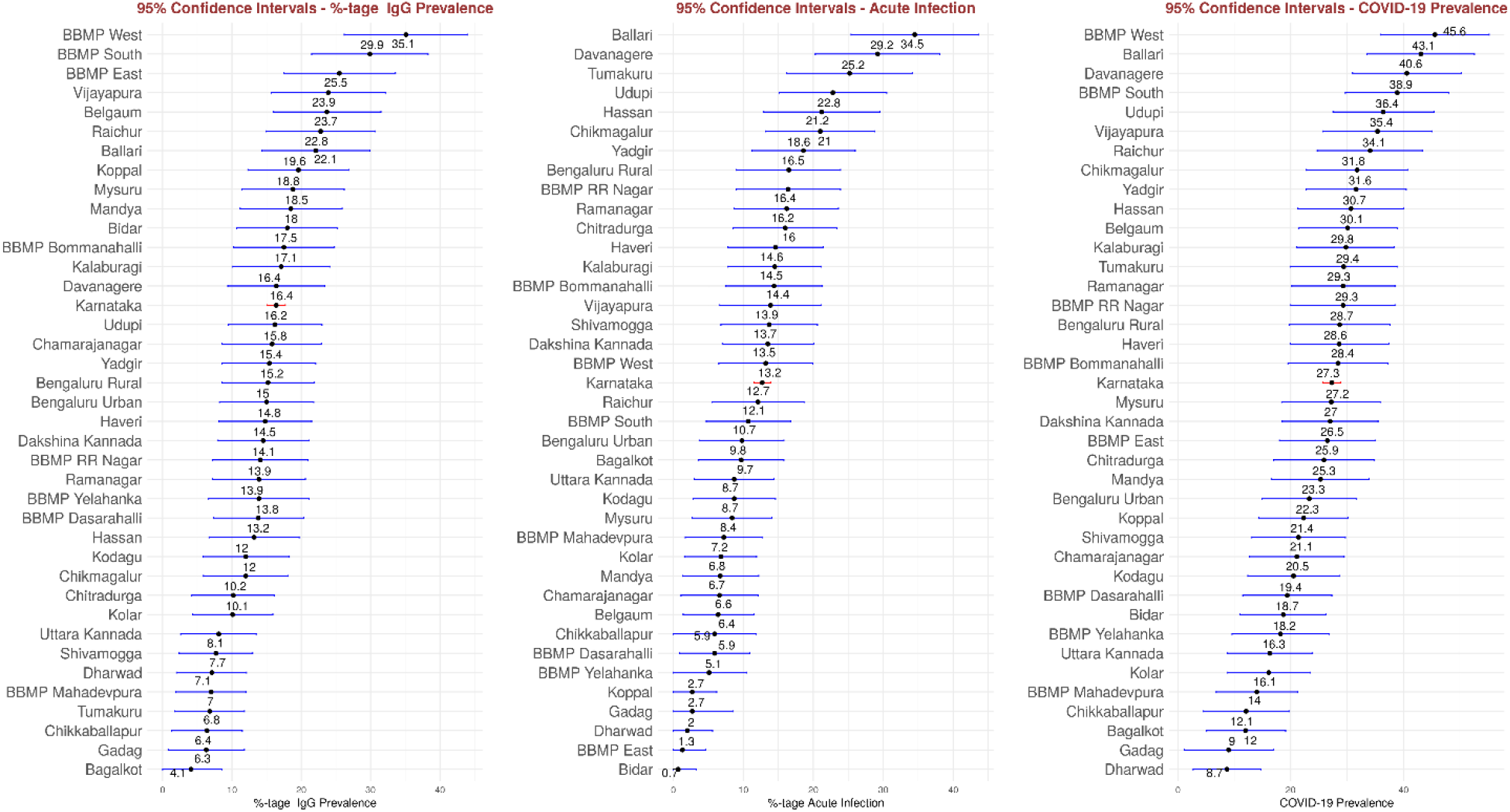
Prevalence (IgG, acute infection, total) and confidence intervals across units. Karnataka is marked in orange.

**Figure 8:**
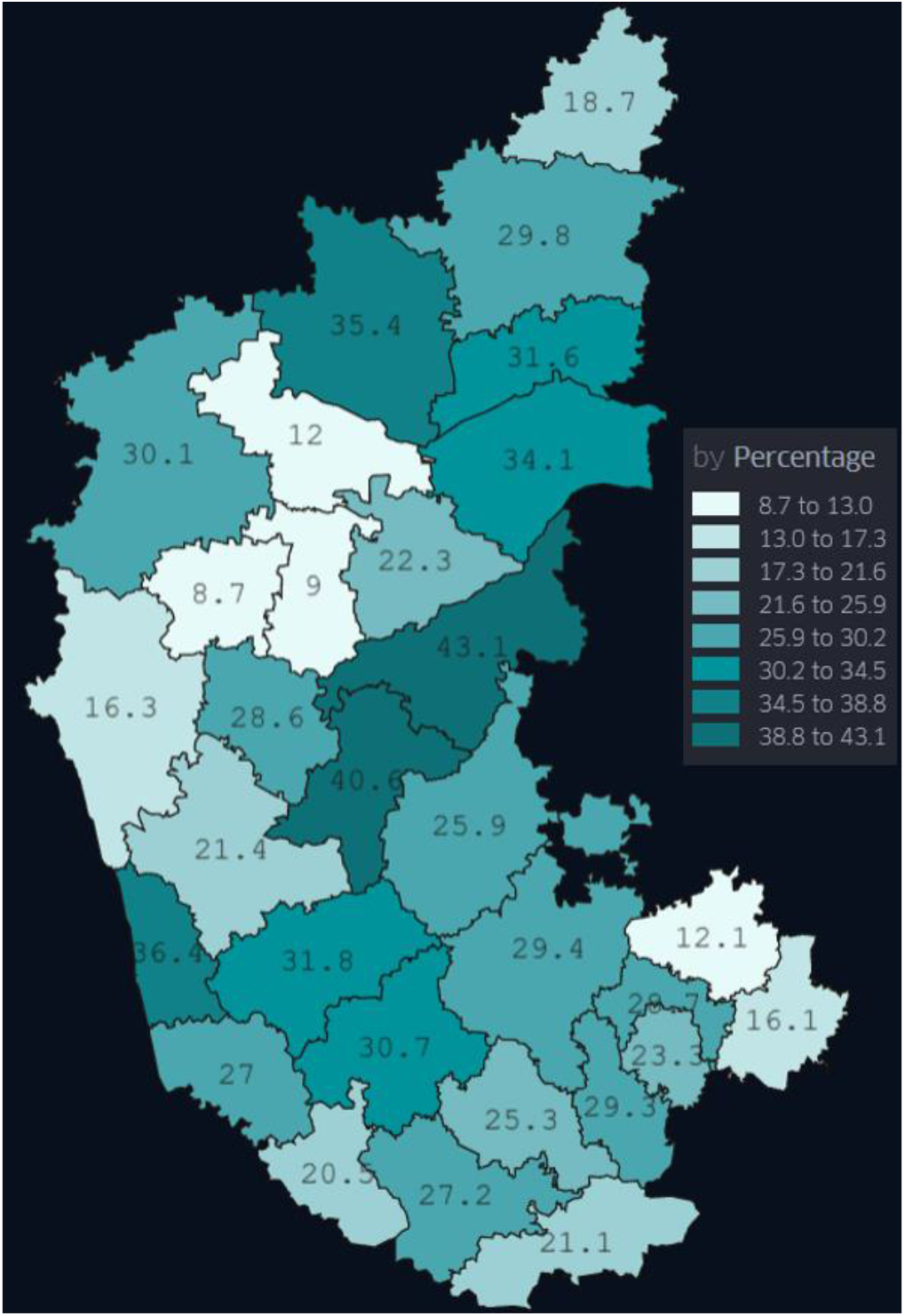
Heat map representing the total prevalence (as percentage of the unit population) across the 30 districts of Karnataka

**Figure 9:**
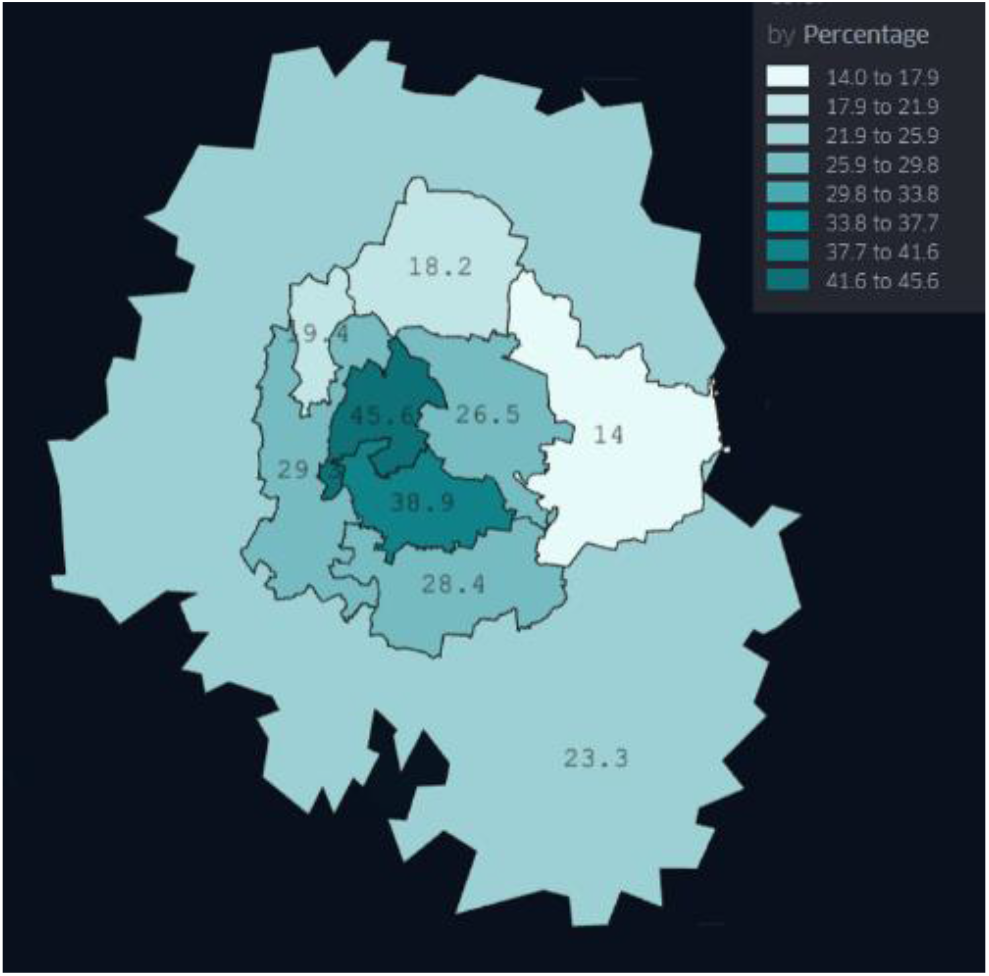
Heat map representing the total prevalence (as percentage of the unit population) across the units of Bengaluru district

### Explanatory variables

A generalized linear model-based multinomial regression indicated that nausea, headache, chest-pain, rhinorrhoea, cough, sore throat, muscle ache, fatigue, chills, and fever are significant variables that predict active infection (p-value < 0 ·05 for a Wald test). Fever is the most significant variable for predicting active infection among the symptoms. Additional variables that predict active infection are attendance at the Out-patient Department (OPD) of the hospitals and contact with COVID-19 positive patients (again with p-value < 0 ·05 for the Wald test). Diarrhea, chest-pain, rhinorrhea, and fever predict the presence of IgG antibodies to some extent, with diarrhea having the highest weight among the three. Additional variables that predict the presence of IgG are professions that involve greater contact with the public (bus conductors or auto drivers, vegetable vendors), residence in containment zones, time since the first 50 cases in the district, and the level of the district’s and the *taluk*’s urbanization (Figure 10 and Table 4).

**TABLE 4:**
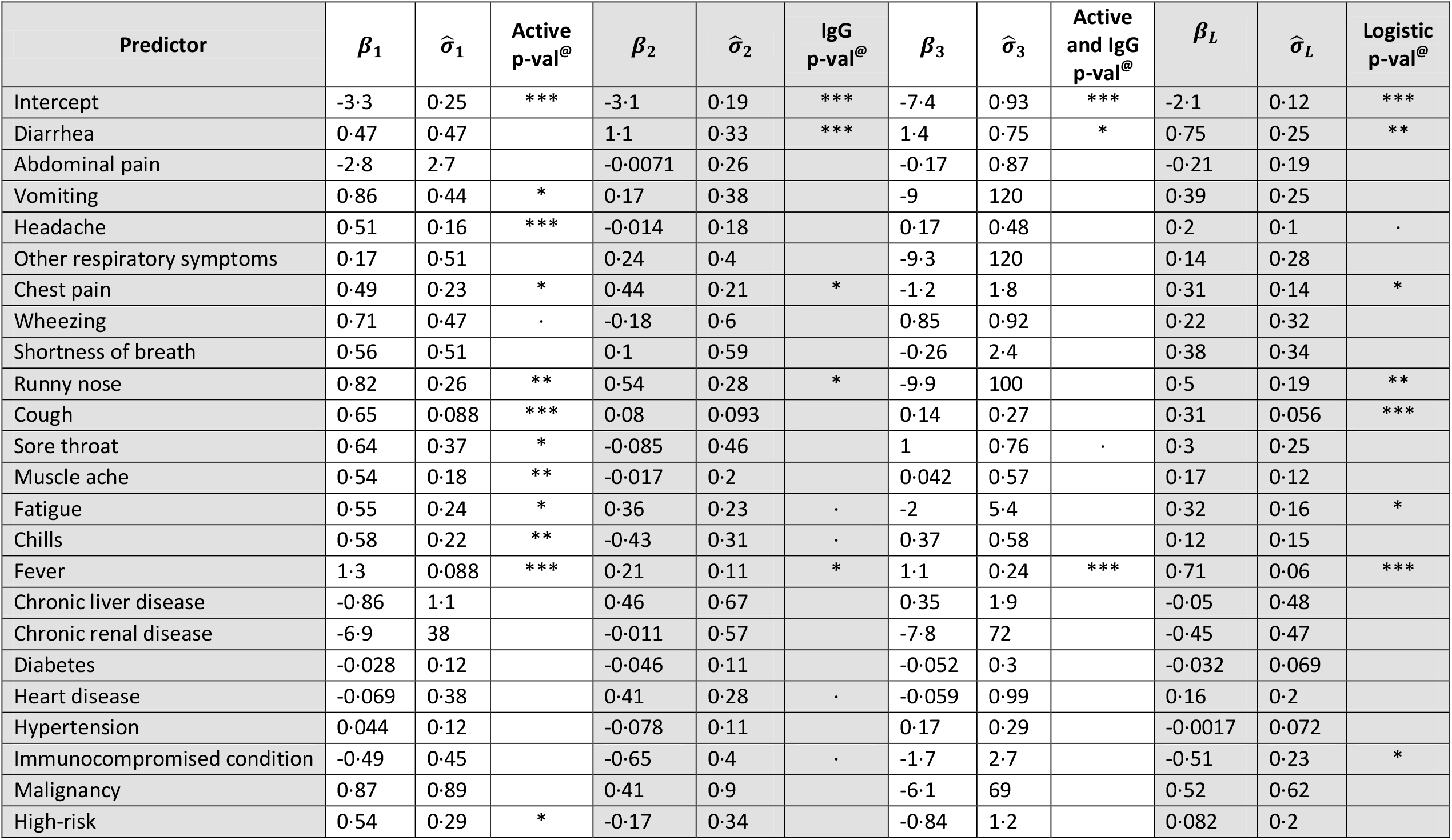
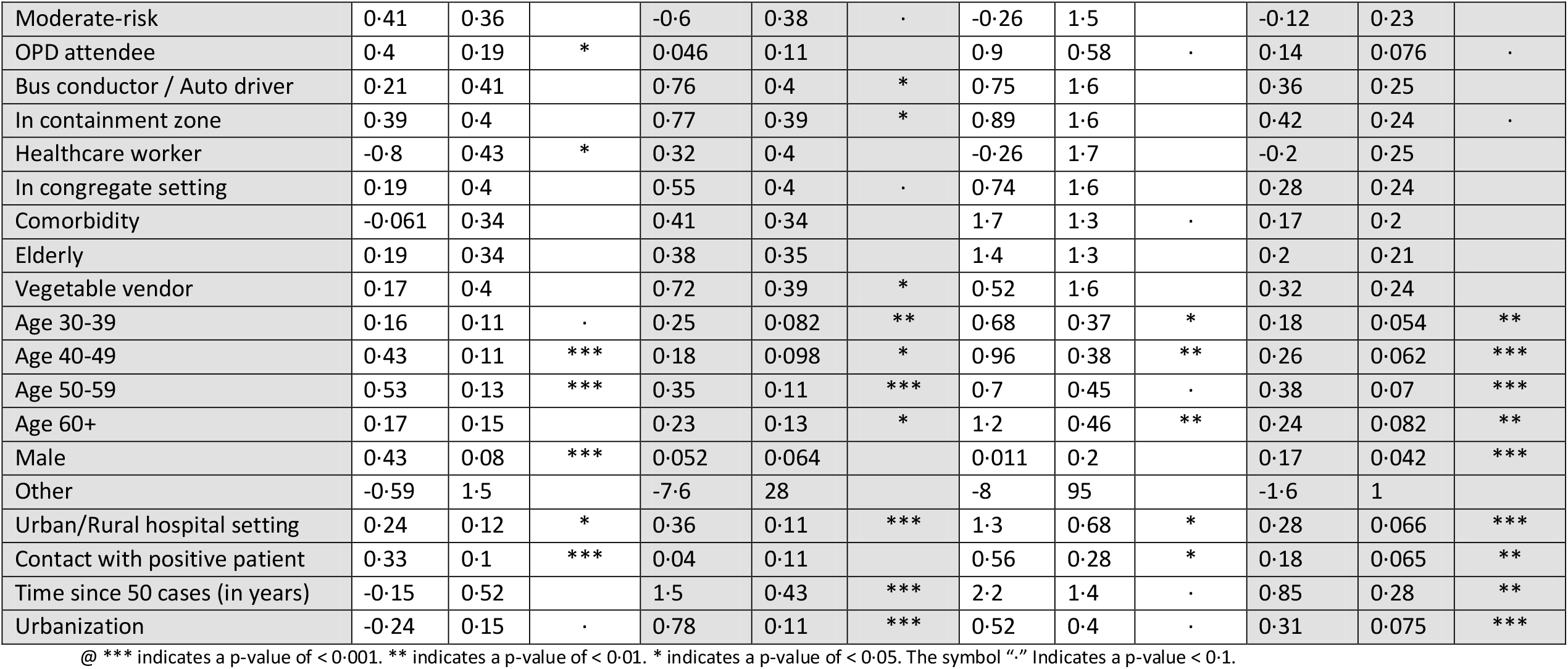
Generalized Linear Model: Prediction of Active, IgG and simultaneous IgG & Active Infection.

**Figure 10:**
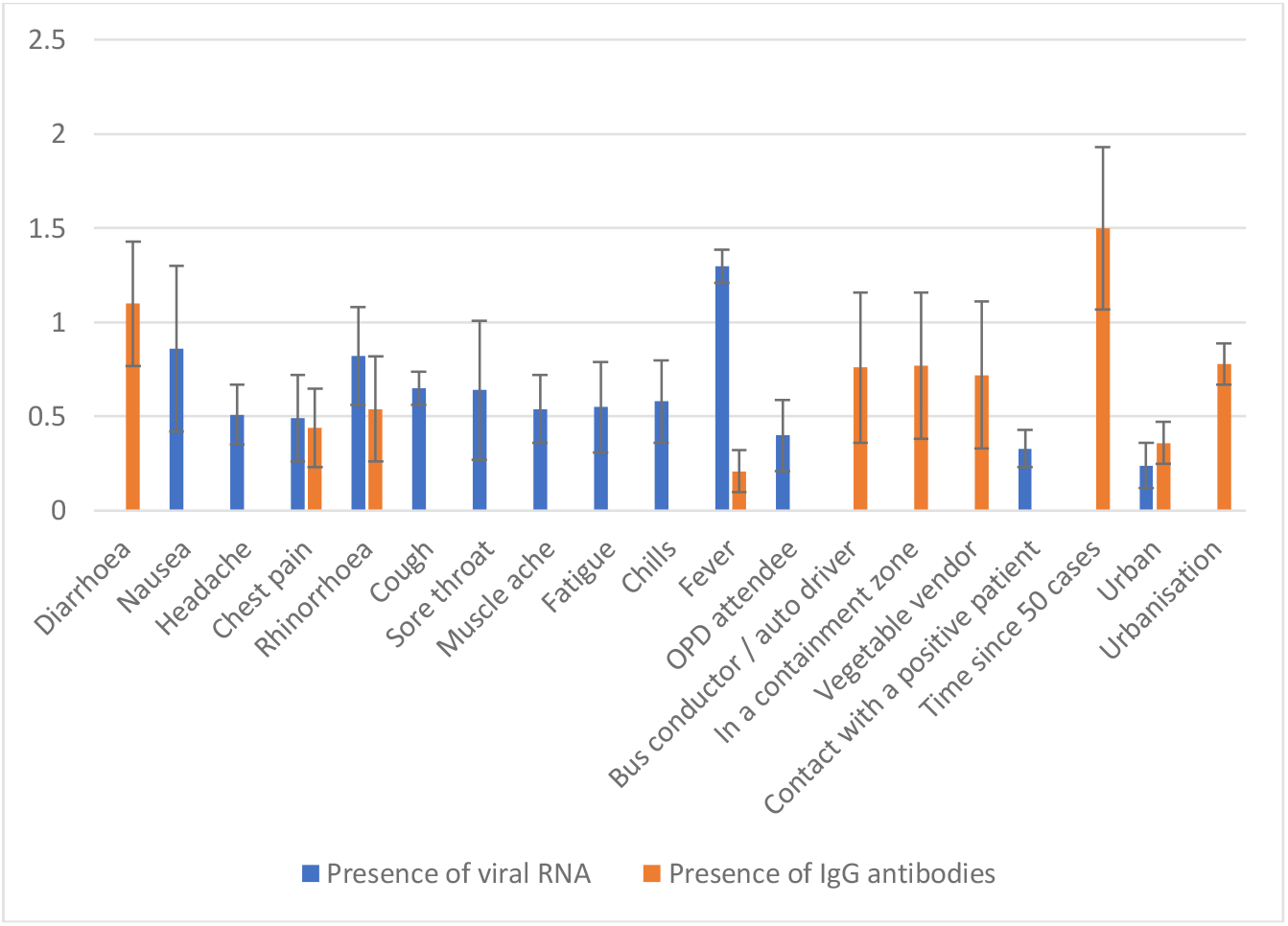
Weights and error bars on a subset of independent variables in a multinomial regression whenever the p-value is lower than 0.05. The abscissa lists the independent variables (symptoms and other factors). The ordinate refers to the weight assigned by the multinomial regression for those selected by the Wald test. The blue bar’s height represents the predictive power for presence of viral RNA. The orange bar is for predicting presence of IgG antibodies. The black error line indicates one standard error.

We found that the RAT is more sensitive on symptomatic individuals since 543 were positive out of 798 RT-PCR-confirmed infected participants with symptoms yielding a sensitivity of 68 ·0% versus 348 being positive out of 742 RT-PCR-confirmed infected participants without symptoms yielding a sensitivity of 46 ·9%.

## DISCUSSION

This is the first study in India, and probably elsewhere, that *jointly estimates* the proportions of people who already had the SARS-CoV-2 Infection (IgG antibody positive) and who currently have an active infection (RT-PCR / RAT positive).

The study has several additional strengths. First, we conducted the study throughout the state of Karnataka using the sentinel sites, thereby leveraging the state’s comprehensive surveillance platform. The sampling frame serves as a reference standard and can be used for population-representative surveillance in the future. Second, we used a serological test for IgG with high sensitivity (0 ·921) and specificity (0 ·977), thereby yielding a better predictive value for a positive test. Lastly, we assessed the prevalence in the key subgroups of populations with differential risk of contracting the SARS-CoV-2 virus.

An estimation of the IgG prevalence alone would have assessed the state’s burden at 16 ·4% prevalence. In contrast, the dual assessment of viral markers and antibodies gave us not only the IgG prevalence but also evidence of active infection of 12 ·7% and a total COVID-19 burden of 27 3%. This significantly larger estimate calls for an entirely different response from the state and highlights our survey’s benefits. We reported that 1 ·8% (95% CI: 1 ·2 – 2 ·3) of the population shows both viral RNA and IgG antibodies. The IgG antibodies form 14-21 days after exposure to the virus, while the RT-PCR test will likely return positive between 7-21 days after exposure. The correlates and implications of the simultaneous presence of viral RNA and IgG antibodies might require further examination in future studies.

Across risk-groups, the elderly and those with comorbidities had a higher prevalence of COVID-19, ^21-23^ suggesting that they are at higher risk of contracting the infection. Despite similar exposure, higher prevalence in them offers the possibility of infection with lower viral dose or that the younger age groups mounted a protective immune response.

The reported IFR due to COVID-19 of 0 ·05% is likely an underestimate, and a function of how well the districts report death data in the state. Studies worldwide found that the IFR of COVID-19 ranged from 0 ·17% to 4 ·16%.^24-26^ The low IFR reported in our study concurs with similar results in India and Asia, including China and Iran.^27-29^ A systematic review of published literature until July 2020 reported the IFR across populations as 0 ·68% (0 ·53%–0 ·82%).^7^ The IFR reported in the survey matches the reported estimates in Mumbai (0 ·05-0 ·10%), Pune (0 ·08%), Delhi (0 ·09%), and Chennai (0 ·13%).^5,12,13,30^ Districts with low CIR suggest that this might be the actual proportion by which we might be missing cases in Karnataka.

Our regression analysis determined which symptoms accurately predict active and past infections. Among the symptoms, we found that diarrhea, chest pain, rhinorrhea, and fever predict the presence of past Infection (IgG antibodies). This suggests that COVID-19 may have consequences that last beyond the active infection period. Diarrhea might suggest that the gastrointestinal tract manifestations might stay longer and might have implications to explore oral vaccines. We also found that ILI symptoms and history of contact with a COVID-19 positive patient can predict active SARS-CoV-2 infection.

The low-risk participants being recruited from hospitals may suggest the existence of a bias in the estimate. The protocol mitigated this by sampling systematically only among pregnant women and attendees of OPD. These participants are likely to have come from afar, thus providing information on prevalence outside the immediate hospital vicinity. The sampling from the congregate settings in the neighborhood of the hospital was made systematic to reduce sampling bias. The elderly and those with comorbidities were to be taken from an elderly list (from the census) and a list (compiled in April 2020) of vulnerable individuals with non-communicable diseases. Any deviation from the protocol would have introduced a bias. Hence, we used a design effect of 3 to account for these factors.

The low sensitivity and cost of RAT consideration led to a survey design in which the low-risk participants were not administered the RAT. Further, due to logistical issues, serum samples from one of the *taluk* hospitals were not available, and the corresponding participants had no antibody test outcomes. Our statistical methodology was designed to handle these issues and make the best use of all the available data.

The progress of the pandemic has been non-uniform, given the wide variation of COVID-19 burden from 8 ·7% to 43 ·1% across various regions of the state. Regions with low IgG prevalence require a targeted public health response. A revision in the testing strategy may be required in districts with high CIR. Given the predictive power of specific symptom complexes for active infection, the state should employ syndromic surveillance to detect active transmission areas. The high sensitivity of the RAT in symptomatic participants indicates that it is better-suited as a point-of-care test when people present themselves with symptoms.

In conclusion, our comprehensive survey and analysis provide insights on the state of the pandemic in the different districts of Karnataka and the varying levels of prevalence across the different stratifications based on age, gender, and risk. We also provide important epidemiological metrics such as IFR, CIR and their variation across geographical regions and population strata (Figure 11). Indeed, establishing district-level facility-based surveillance to systematically monitor the trend of Infection in the long term to inform local decision-making at the district level would facilitate and augment the necessary public health response towards the COVID-19 epidemic in Karnataka. It also helps identify regions with high severity of the disease, identify at-risk populations, and enable evidence-based intervention and resource allocation to manage the pandemic effectively. Repetition of the survey can better inform changes in the extent and speed of transmission and help evaluate the potential impact of containment strategies over time in different parts of the state. Above all, this study’s findings hold significant potential to improve clinical management and guide public health interventions to reduce the burden of COVID-19 in India and other lower and middle-income countries.

**Figure 11:**
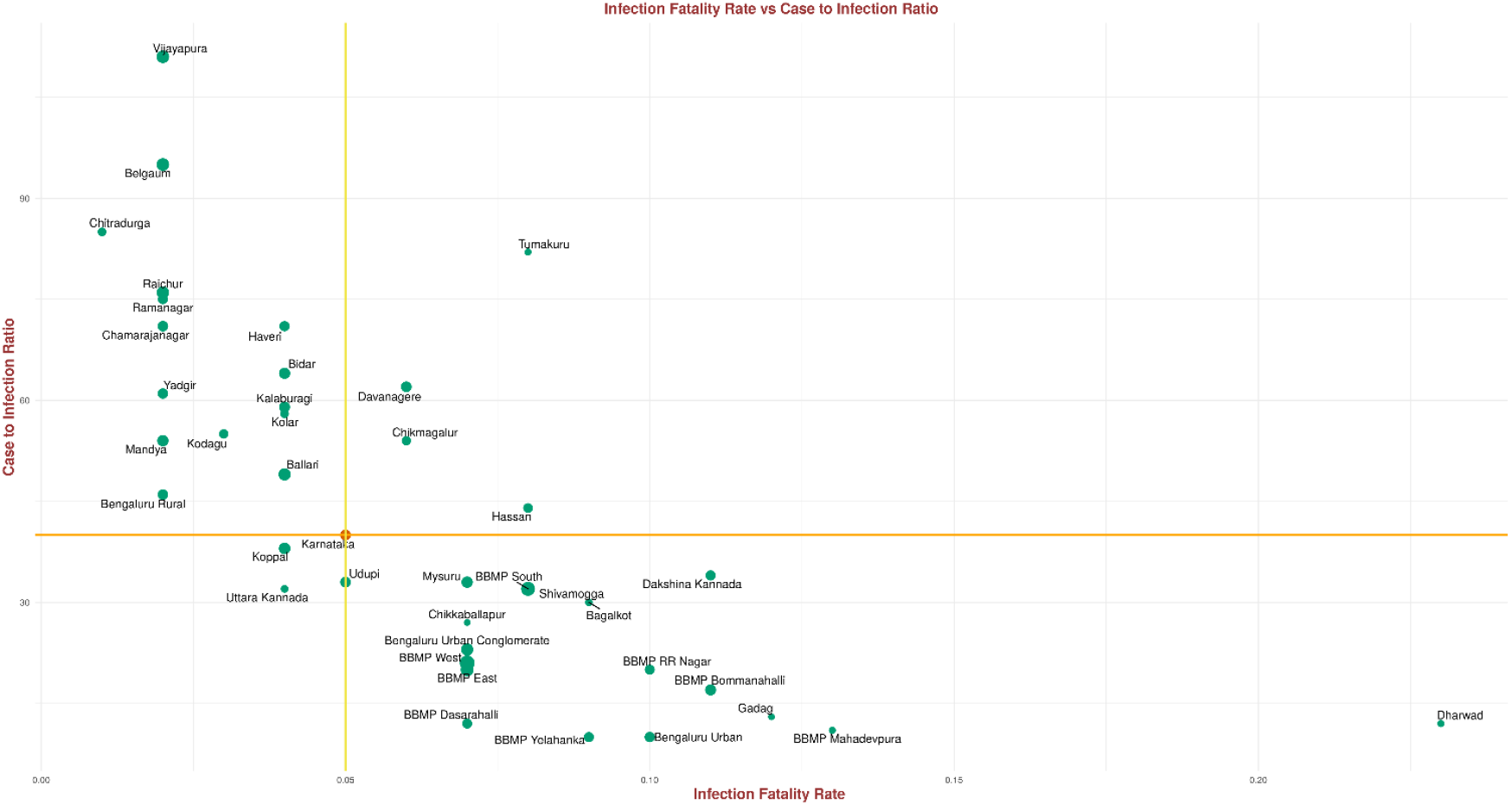
Scatter plot of CIR versus IFR. The size of the point indicates the IgG prevalence in the units. The horizontal and the vertical lines intersect at Karnataka’s IFR and CIR. We move clock-wise from the upper-left quadrant. A unit in the upper-left quadrant with a larger green disk has high IgG antibody prevalence, low IFR, and high CIR. Such a unit is missing cases and deaths. A unit with a larger green disk in the upper-right quadrant has high IgG antibody prevalence, high IFR, and high CIR. Such a unit is also likely missing cases, but death reporting is better than average. A unit with a larger green disk in the bottom-right quadrant has high IgG antibody prevalence, high IFR, and low CIR. Such a unit has done well in identifying cases and has better-than-average reporting of deaths. A unit with a larger green disk in the bottom left has low IFR and low CIR. Such a unit has seen a surge in cases, but has done well in identifying cases, and has low fatality rates perhaps due to good clinical practices that could be studied and replicated elsewhere.

## CONTRIBUTORS

The survey was a collaborative effort of the Department of Health and Family Welfare, National Institute of Mental Health and Neuro-Sciences, Indian Institute of Public Health - Bangalore, Indian Institute of Science, Indian Statistical Institute (Bangalore Centre), UNICEF, MS Ramaiah Medical College, Bangalore Medical College, and others. The protocol was designed by Prof Giridhara R. Babu and his team at the IIPH, Bangalore, along with the following members of the Technical Advisory Committee – Dr. Lalitha R, Dr. Lalitha K, and Dr. Pradeep B. The Technical Advisory Committee chaired by Prof Dr. M. K. Sudarshan reviewed and provided feedback on the design and implementation of the survey. Dr. M. R. Padma, Dr. Mohammed Shariff, under the supervision of Dr. Parimala Maroor, Project Director IDSP, coordinated the implementation at the state level. The technical review group chaired by the Director, DHFWS, approved the conducting of the study. Mr. Jawaid Akhtar, Mr.Pankaj Kumar Pandey reviewed the protocol, led the implementation and were involved in writing and reviewing the manuscript. Professors Giridhara R. Babu, Siva Athreya, and Rajesh Sundaresan planned and executed the data analysis, arrived at the findings, and wrote the first draft and revisions of the manuscript. All authors reviewed and approved the final manuscript.

## DATA SHARING

The data are accessible to researchers upon formal request for data addressed to the Commissioner, Health and Family Welfare Services, Government of Karnataka.

## Supporting information

Sero_survey__Manuscript_Supplementary file_Medrxiv.pdf

## Acknowledgments

We would like to express our thanks to: Dr Arundathi, IAS, MD – NHM, Dr. Patil Om Prakash R Director – DHFWS, Dr. Prakash, State nodal officer for COVID19 and State Surveillance Unit for their support; DSOs, DAPCU officers, AMOs & Medical officers, District Microbiologists and District Epidemiologists for coordinating and implementing survey and providing guidance for sample collection as per sample size to health facility lab staff and coordinating for sample transportation to mapped RT-PCR & antibody testing ICMR labs; Lab Nodal Officer and staff of ICMR labs for RT-PCR testing and IgG antibody testing; Mr. Ramesh and team for providing a robust web platform for data collection; Lab technicians, Counsellor –ICTC & NCDC, Staff Nurse, Health workers for filling data in the survey App, collection of samples and sending samples to higher labs; Ms. Manjushree, Entomologist, DHFWS for helping in fetching RAT/RT-PCR results from ICMR Portal, Ms. Maithili Karthik and Ms. Sindhu ND, PHFI, for help with the line list matching; Mr. Nihesh Rathod, Indian Institute of Science, for the generation of Karnataka and Bengaluru Urban Conglomerate maps; Nitya Gadhiwala and Abhiti Mishra of the Indian Statistical Institute for help in collation of COVID-19 data from Karnataka state bulletins and R graphics; All the study participant for providing their consent to be part of this survey.

