## Supplementary material for "The burden of active infection and anti-SARS-CoV-2 IgG antibodies in the general population: Results from a statewide survey in Karnataka, India": Sero_survey__Manuscript_Supplementary file_Medrxiv.pdf

### APPENDIX A

**SUPPLEMENTARY TABLE I: Seroprevalence of IgG antibodies against SARS-CoV2 and active infection in Bengaluru UrbanConglomerate (*N* = 3617)**

| BBMP Zones | Samples[ψ] | %- IgG againstSARS-CoV2[†] | %-Active Infection[†] | %-Prevalence of COVID-19[†] |
|---|---|---|---|---|
| Bengaluru Urban Conglomerate | 3617 | 22 (19·1–24·9) | 9·2 (7·1–11·3) | 29·8 (26·5–33) |
| BBMP West | 405 | 35·1 (26·2–44) | 13·2 (6·5–19·9) | 45·6 (36–55·2) |
| BBMP South | 422 | 29·9 (21·5–38·3) | 10·7 (4·7–16·8) | 38·9 (29·7–48·1) |
| BBMP RR Nagar | 381 | 14·1 (7·2–21) | 16·4 (9–23·9) | 29·3 (20–38·5) |
| BBMP Bommanahalli | 394 | 17·5 (10·2–24·8) | 14·4 (7·5–21·3) | 28·4 (19·5–37·3) |
| BBMP East | 416 | 25·5 (17·5–33·6) | 1·3 (0–4·6) | 26·5 (18–35) |
| Bengaluru Urban | 406 | 15 (8·2–21·8) | 9·8 (3·7–15·8) | 23·3 (14·9–31·7) |
| BBMP Dasarahalli | 421 | 13·8 (7·3–20·4) | 5·9 (0·9–10·9) | 19·4 (11·5–27·3) |
| BBMP Yelahanka | 339 | 13·9 (6·6–21·1) | 5·1 (0–10·5) | 18·2 (9·6–26·8) |
| BBMP Mahadevpura | 433 | 7 (1·9–12) | 7·2 (1·7–12·7) | 14 (6·8–21·2) |

ψ Includes only samples that have been mapped to individuals

† Adjusted for sensitivities and specificities of RAT, RT-PCR, and antibody testing kits and procedures

#### APPENDIX B

#### STATISTICAL MODELLING FRAMEWORK

In order to get the prevalence estimates, the following modeling approach was taken. An individual might be in one of four different states:

- $s = 1$: active infection but no IgG antibodies;
- $s = 2$: IgG antibodies present but no evidence of active infection;
- $s = 3$: simultaneous presence of active infection and IgG antibodies;
- $s = 4$: neither active infection nor IgG antibodies.

An individual in state $s = 1$ will nominally have a positive outcome on the RAT and RTPCR tests and a negative outcome on the IgG antibody test. An individual in state $s = 2$ will nominally have a negative outcome on the RAT and the RTPCR tests and a positive outcome on the IgG antibody test. An individual in state $s = 3$ will nominally come out positive on all three tests; we anticipate that this is a small fraction. Finally, an individual in state $s = 4$ will nominally come out negative on all three tests. The third state $s = 3$ recognizes that the viral RNA may persist for some time even after recovery. See Table II of this supplement.

**SUPPLEMENTARY TABLE II: Table of states and nominal test responses $M(s,j)$**

| State $s$ | State description | RAT $j = 1$ | RT-PCR $j = 2$ | IgG $j = 3$ |
|---|---|---|---|---|
| $s = 1$ | Active infection but no IgG antibodies | 1 | 1 | 0 |
| $s = 2$ | IgG antibodies present but no evidence of active infection | 0 | 0 | 1 |
| $s = 3$ | Simultaneous presence of active infection and IgG antibodies | 1 | 1 | 1 |
| $s = 4$ | Neither active infection nor IgG antibodies | 0 | 0 | 0 |

Within each state/unit/stratification, let the fraction of individuals in state $i$ be $p_i$, where $p_i \geq 0$, $i = 1,2,3,4$, and $p_1 + p_2 + p_3 + p_4 = 1$. The tests have imperfect but known sensitivities and specificities, and the test outcomes provide noisy information about the hidden state of an individual. Within each district/unit/stratification, a maximum likelihood estimation procedure then estimates the fraction of individuals $p_1, p_2, p_3, p_4$ in each state. The fraction with active infection is then $p_1 + p_3$, the fraction with IgG antibodies is $p_2 + p_3$, and the total prevalence in the unit (which is the fraction with IgG + active infection) is $p_1 + p_2 + p_3$. This last fraction is likely an underestimate of the disease burden in the unit because our protocol does not allow estimation of the fraction with past infection but recovered by mounting a T-cell immune response.

If the only test that is conducted is the IgG antibody test, then the maximum likelihood estimate of $p_2 + p_3$, adjusted for the sensitivity and the specificity of the IgG test, is the familiar Rogan-Gladen formula: [20]

$$\left[\frac{\widehat{IgG} + specificity(IgG) - 1}{sensitivity(IgG) + specificity(IgG) - 1}\right]_0^1$$

where $\widehat{IgG}$ is the crude estimate of the IgG prevalence in the population (fraction who come out IgG positive among the participants), specificity(IgG) and sensitivity(IgG) are the specificity and the sensitivity of the antibody test, and $[x]_0^1$ is the projection of a real number $x$ to the interval $[0,1]$. Our maximum likelihood procedure, elaborated below, handles the situation when the combination of tests varies across individuals. We can therefore view our approach as an extension of the Rogan-Gladen formula. See for a simple situation when only one hidden state probability is to be estimated, but multiple tests are conducted on each participant.

We computed odds ratios by focusing on the relevant subcategories: male/female or other/female with the female category taken as reference; high-risk/low-risk, moderate-risk/low-risk with the low-risk category taken as reference; etc. Important independent variables are identified using multinomial regression and a generalized linear model with a custom link function accounting for the sensitivities and the specificities of the tests. This is elaborated in the rest of this subsection. Confidence intervals are computed using the Fisher information (for prevalence estimation or odds ratio computation) or using the observation information (when identifying important independent variables).

We now elaborate on the statistical methodology. The indices $j = 1, 2$, and 3 will stand for RAT, RT-PCR andIgG antibody tests, respectively. Let $t = (t_1, t_2, t_3)$ denote the test pattern: if $t_j = 1$, then the test $j$ had a valid outcome, else test $t_j = 0$, and the test $j$ was either not conducted or had an invalid outcome. Let $y = (y_1, y_2, y_3)$, where $y_j \in \{0,1, NA\}$; these three values stand for a negative outcome, a positive outcome, or an invalid outcome, respectively. Consequently $y_j = 0$ or 1 when $t_j = 1$ (a valid test outcome), and $y_j = NA$ when $t_j = 0$ (an invalid test outcome or the test was not conducted).

An individual in state $s = 1$ will have a nominal positive RAT test outcome, a nominal RT-PCR test outcome, and a nominal negative IgG test outcome. Nominal outcomes can be similarly written for the other states. Write $M\,(s, j)$ for the nominal test outcome for an individualin state $s$; $M\,(s, j) = 1$ indicates a nominal positive outcome, and $M\,(s, j) = 0$ indicates a nominal negative outcome. The last three columns of Table II of the supplement provide the nominal responses on the RAT, the RT-PCR and the IgG antibody tests for each of the four states.

The actual test outcomes may however differ from the nominal outcomes. Let $\sigma(0, j)$ denote the specificity of test $j$, and let $\sigma(1, j)$ denote the sensitivity of test $j$. The specificities and sensitivities used for the estimations are as given in Table III of this supplement.

**SUPPLEMENTARY TABLE III: The sensitivities and specificities**

| $\boldsymbol{\sigma(m, j)}$ | RAT $\boldsymbol{j = 1}$ | RT-PCR $\boldsymbol{j = 2}$ | IgG $\boldsymbol{j = 3}$ |
|---|---|---|---|
| Specificities ($m$ = 0) | 0·975 | 0·97 | 0·977 |
| Sensitivities ($m$ = 1) | 0·5 | 0·95 | 0·921 |

To describe the probability of test outcomes, we first consider an example. Take an individual in state $s = 1$, test pattern $t = (1,1,1)$, and test outcomes $y = (0,1,0)$. This is a situation where RAT is false negative and will therefore involve one minus the sensitivity of the RAT test, the RT-PCR test is true positive and will involve the sensitivity of the RT-PCR test, and the IgG antibody test is true negative and will involve the IgG test's specificity. Thus, the conditional probability of the test outcome, given the state and the test pattern, is

$$q(y = (0,1,0) | s = 1, t = (1,1,1)) = [1 - \sigma(1,1)] \cdot [\sigma(1,2)] \cdot [\sigma(0,3)]$$

In general, for an individual in state $s$ with test pattern $t = (t_1, t_2, t_3)$, the conditional probability of test outcome $y = (y_1, y_2, y_3)$ is given by

$$q(y|s, t) = \prod_{j:t_j=1} \sigma(M(s, j), j)^{\mathbf{1}\{M(s,j)=y_j\}} \cdot [1 - \sigma(M(s, j), j)]^{1-\mathbf{1}\{M(s,j)=y_j\}}$$

where $M$ and $\sigma$ are given in Supplementary Tables II and III, respectively, and $\mathbf{1}\{M(s,j) = y_j\}$ is the characteristic function or indicator function that tests the condition $M(s,j) = y_j$, i.e., it is 1 if $M(s,j) = y_j$ holds and it is 0 otherwise.

There are $N$ participants. Each participant has a list of attributes denoted $x(n)$ (e.g., district, age, gender, risk category, risk sub-category, etc.). Each participant also has a hidden state $s(n)$, a test type $t(n)$, and a test outcome $y(n)$. Let the probabilities of an individual being in the four states be $p = (p_1, p_2, p_3, p_4)$, with $p_s \geq 0, \sum_s p_s = 1$. The probability that an individual $n$'s test outcomes are $y(n)$ given test pattern $t(n)$ is:

$$P\big(y(n)\big|t(n)\big) = \sum_{s=1}^{4} P\big(y(n), s\big|t(n)\big) = \sum_{s=1}^{4} p_s \cdot q(y(n)|s, t(n))$$

where $q(y|s,t)$ is as given above. We have thus arrived at a parametric model with a vector parameter $p$.

1. *Maximum likelihood estimation.* Let us focus on a stratum, say district $D$ with $N_D$ participants. Denoting $x(D)$, $t(D)$, and $y(D)$ to be the restriction of the data to the stratum $D$, assuming independence across individuals, in this stratum, we arrive at the likelihood:

$$L\big(p; x(D), t(D), y(D)\big) \coloneqq \prod_{n:n\in D} \left(\sum_{s=1}^{4} p_s q(y(n)|s, t(n))\right)$$

- The mapping $p \mapsto \log L(p; x(D), t(D), y(D))$ is concave, so a numerical, iterative, gradient descent procedure is used to arrive at the maximum likelihood estimate for $p$ in this stratum, denoted by $\hat{p}(D) = (\hat{p}_1(D), \hat{p}_2(D), \hat{p}_3(D))$. The estimate of active prevalence in the stratum is $\hat{p}_1(D) + \hat{p}_3(D)$, the estimate of IgG prevalence in the stratum is $\hat{p}_2(D) + \hat{p}_3(D)$, and the estimate of the total prevalence (past and active infection) in the stratum is $\hat{\wp}(D) \coloneqq \hat{p}_1(D) + \hat{p}_2(D) + \hat{p}_3(D)$. This provides the joint estimation of IgG prevalence, active infection prevalence, and total prevalence of COVID-19 (IgG and active prevalences).

- The confidence interval estimations come from the Fisher information matrix associated with the aforementioned parametric model and a design effect of 3. Let us elaborate. For protocol and logistical reasons already explained earlier, the test patterns differ across the participants. Consider a fixed test pattern and the associated Fisher information matrix for one sample coming from this test pattern. The overall per-sample Fisher information matrix for the collection is an average with weights in proportion to the observed test patterns. In other words, the overall per-sample Fisher information matrix may be written as

$$I = \sum_{t} w_t I(t)$$

  where $w_t$ is the fraction of individuals in the stratum whose test pattern is $t$ and $I(t)$ is the Fisher information matrix of the parametric model $P(y|t)$ with test pattern $t$ and parameter vector $p$. If there were enough participants with pattern $t$ and if we used only test outcomes from this pattern, in the large sample size asymptotic, we can invoke asymptotic normality – the covariance of the estimated vector parameter (after re-scaling by square-root of the number of samples of pattern $t$) will approach $I(t)^{-1}$. Given that we have multiple test patterns, the covariance of the estimated vector parameter $\hat{p}(D)$ can be taken to be approximately $v(\hat{p}(D)) \coloneqq \frac{I^{-1}}{\sqrt{N_D}}$ . One may view $I$ as a *hybrid* observation/Fisher information matrix. The estimations of IgG, active prevalence, and total prevalence being linear combinations of $p_1$, $p_2$ , and $p_3$, their estimated variances are given by $\frac{u^T I^{-1} u}{\sqrt{N_D}}$, where $u$ is either $(0,1,1)^T$ (IgG prevalence) or $(1,0,1)^T$ (prevalence of active infection) or $(1,1,1)^T$ (for total COVID-19 prevalence), and the superscript denotes matrix transposition.

2. *State-wide estimates*. The Karnataka estimates are obtained after weighting for district populations:

$$\hat{p} = \sum_{D} w(D)\, \hat{p}(D)$$

   where $w(D)$ is the fraction of Karnataka population that lives in district $D$. Assuming independence across districts, the covariance is:

$$\sum_{D} w(D)^2 cov(\hat{p}(D))$$

The weighted estimates and confidence intervals of IgG, active, and total COVID-19 prevalence follow the same procedure as outlined.

3. *Odds ratios*. Let us write the odds ratio for an example. Let $D_H$ and $D_L$ denote the high-risk category and the low-risk category stratifications, respectively. The odds of being infected given high-risk versus the odds of being infected given low-risk is:

$$\frac{odds(D_H)}{odds(D_L)} = \frac{\hat{\wp}(D_H)/[1-\hat{\wp}(D_H)]}{\hat{\wp}(D_L)/[1-\hat{\wp}(D_L)]}$$

The 95% confidence intervals for prevalence $\hat{\wp}(D_H)$ and $\hat{\wp}(D_L)$ automatically yield 90% confidence intervals for the odds ratios.

4. *Important covariates*. To identify which features are important predictors of active infection, IgG, etc., we introduce a multinomial logit function for the probability of three of the four hidden states. The multinomial logit function is taken to be an affine function of the independent variables, with the parameters of the affine function being the weights. That is,

$$p_s \propto exp\{\beta_{s,0} + \beta_s^T x\}, \quad s = 1,2,3$$

where $x$ is the vector of independent or explanatory variables. All categorical variables were encoded using the *one-hot* encoding method and the references (that were used in the odds-ratio calculations) were removed. The scalar $\beta_{s,0}$ is the so-called intercept. The vector $\beta_s$ denotes the weights for the independent variables in $x$. The resulting mapping

$$\beta = ((\beta_{s,0}\, \beta_s), s = 1,2,3) \mapsto L(p(\beta);\, x(D), t(D), y(D)),$$

in general, is not concave. The iterative schemes will therefore only take us to a local maximum. We take the best solution after multiple restarts of this iterative procedure. The inverse of the observation information matrix at the searched local maximum gives the covariance of $\beta$. The standard Wald's test then identifies the important factors. We compare the outcome with a similar outcome for the much simpler logistic regression where the target is taken to be $y_1 \oplus y_2 \oplus y_3$, which reports positive if any of the RAT, RTPCR or IgG antibody test result is positive.

The computations were done in R version 4.0.3. The optimizations employed the optim and the constrOptim functions with the quasi-Newton BFGS method.

We now present some interesting aspects of the proposed statistical methodology.

1) Consider an individual whose RAT outcome is negative, RT- PCR test outcome is positive, and IgG outcome is negative. A statistical inference for active prevalence that declares a participant as having active infection when either the RAT or the RTPCR outcome is positive, but ignores the individual's IgG test outcome, will suffer from the following issues.

   - It will have a higher sensitivity at the expense of a poorer specificity, and therefore will result in more false positives.

   - It also does not exploit the enhanced evidence for active prevalence arising from the lack of IgG antibodies.

   Similarly, a statistical inference for IgG prevalence that ignores the RAT and RT-PCR outcomes does not recognize that a positive RT-PCR test may enhance evidence of lack of IgG antibodies.

   It is therefore not sufficient to look only at IgG test outcomes for estimating IgG prevalence; similarly, it is not sufficient to look only at RAT + RT-PCR outcomes for estimating active prevalence. A combined estimation, that makes the best use of all the available data, is therefore essential.

2) The test patterns were not the same across participants.

   - The low sensitivity of the RAT test and cost consideration led to a protocol design in which the low-risk category participants were not administered the RAT test.

   - The ICMR protocol required that if the RAT outcome is positive, the usual protocol for positive cases ensues, and the RT-PCR test will not be conducted.

- The RT-PCR results were not retrievable in time for roughly 6% of the participants.

- Due to logistical issues, serum samples from one of the taluk hospitals were not available, and the corresponding participants had no antibody test outcomes.

The evidence of active and IgG prevalence coming from the test outcomes of a participant for whom all three tests were done should be weighted differently from those for whom only a subset of the test outcomes is available.

3) The RAT, RT-PCR, and the IgG antibody tests are nonideal. Their measured sensitivities and specificities should be appropriately used to adjust the prevalence estimates.

Our proposed four-state model permits an expression of conditional probabilities, given a test pattern, in terms of the hidden state's probabilities. The resulting parametric model has a concave log-likelihood function and is amenable to maximum-likelihood estimation using a numerical procedure. As already highlighted, the methodology generalizes the Rogan-Gladen formula[20] to situations with multiple hidden states, noisy observations, and differing test patterns across individuals.

The four-state model also enables us to highlight the importance of symptoms in predicting active infection and in recognizing that the gastro-intestinal tract may be involved in the disease. A naïve logistic regression on the variables for the target $y = y_1 \oplus y_2 \oplus y_3$, a crude indicator of either active infection or presence of IgG, leaves out some symptoms, picks diarrhea with reduced weight, does not recognize that certain symptoms or contact with a COVID-19 positive patient as important for predicting active infection, and does not connect diarrhea with past infection.

It should be noted that the multinomial regression to assess importance of variables involves a non-concave maximization (in the analysis of important features), and our iterative scheme may have settled at a local maximum. But we are confident that this is not the case since the data analysis is in-line with clinical findings (symptoms predicting active infection).

#### APPENDIX C

#### FLOWCHART SHOWING VALID SAMPLES INDICATING HOW WE ARRIVED AT OUR LINE LIST.

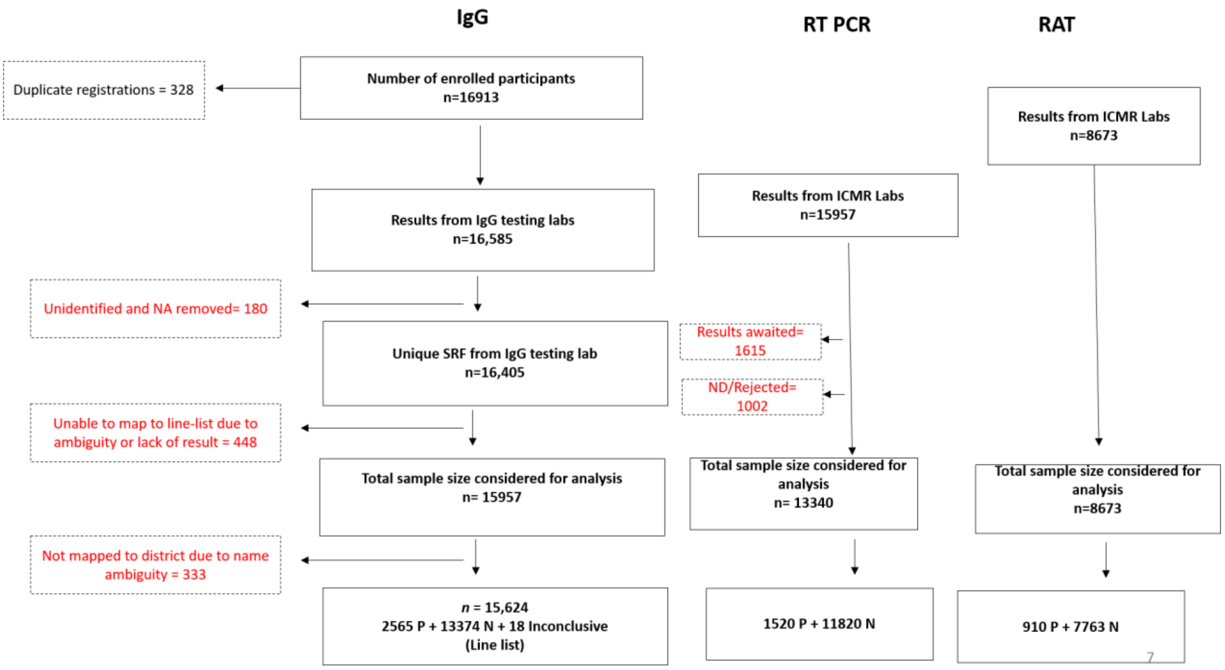


Supplementary Figure I: Flow chart showing valid samples indicating how we arrived at our line list.

The flowchart above shows how we arrived at our line list based on the three different tests conducted and the number of participants.

#### APPENDIX D

#### DESCRIPTION OF UNITS AND HEALTH FACILITIES

Each district of Karnataka was taken to be a unit for the survey but in case of Bengaluru, it was further subdivided into nine units. Hence there will be a total of 38 units. See the description of sampling units in Supplementary Table IV. From each unit, between five and fifteen subdistricts or *taluks* were selected. In each taluk, either the district hospital or the taluk hospital or the primary/community health facility was chosen to conduct the survey. The locations of the facilities shown in Figure 1 .

##### SUPPLEMENTARY TABLE IV: Description of the sampling units

| S. No. | Unit | Area (km$^2$) | Estimated population (2020) |
|---|---|---|---|
| 1 | Bagalkot | 6575 | 2141784 |
| 2 | Ballari | 8450 | 3096512 |
| 3 | Belgaum | 13415 | 5354714 |
| 4 | Bengaluru Rural | 2259 | 1139694 |
| 5 | Bengaluru: BBMP Bommanahalli | 98 | 1104157 |
| 6 | Bengaluru: BBMP Dasarahalli | 28 | 536362 |
| 7 | Bengaluru: BBMP East | 92 | 2030933 |
| 8 | Bengaluru: BBMP Mahadevpura | 172 | 1073525 |
| 9 | Bengaluru: BBMP RR Nagar | 110 | 895343 |
| 10 | Bengaluru: BBMP South | 72 | 2345502 |
| 11 | Bengaluru: BBMP West | 39 | 1524786 |
| 12 | Bengaluru: BBMP Yelahanka | 99 | 660342 |
| 13 | Bengaluru: (rest of) Bengaluru Urban | 1481 | 1076763 |
| 14 | Bidar | 5448 | 1903546 |
| 15 | Chamarajanagar | 5101 | 1076085 |
| 16 | Chikkaballapur | 4524 | 1364310 |
| 17 | Chikmagalur | 7201 | 1136558 |
| 18 | Chitradurga | 8440 | 1803209 |
| 19 | Dakshina Kannada | 4560 | 2335183 |
| 20 | Davanagere | 5924 | 2102415 |
| 21 | Dharwad | 4260 | 2099469 |
| 22 | Gadag | 4656 | 1157112 |
| 23 | Hassan | 6814 | 1840223 |
| 24 | Haveri | 4823 | 1759087 |
| 25 | Kalaburagi | 10951 | 2976434 |
| 26 | Kodagu | 4102 | 560797 |
| 27 | Kolar | 3969 | 1695959 |
| 28 | Koppal | 7189 | 1594147 |
| 29 | Mandya | 4961 | 1851525 |
| 30 | Mysuru | 6854 | 3374961 |
| 31 | Raichur | 6827 | 2187486 |
| 32 | Ramanagar | 3556 | 1140574 |
| 33 | Shivamogga | 8477 | 1864370 |
| 34 | Tumakuru | 10597 | 2784099 |
| 35 | Udupi | 3880 | 1307243 |
| 36 | Uttara Kannada | 10291 | 1516250 |
| 37 | Vijayapura | 10494 | 2571844 |
| 38 | Yadgir | 5213 | 1412445 |
